# Charting Brain Structure in 22q11.2 Deletion Syndrome with Clinical Neuroimaging

**DOI:** 10.1101/2025.11.25.25339898

**Authors:** Benjamin Jung, J. Eric Schmitt, Jakob Seidlitz, Jenna M. Schabdach, Shivaram Karandikar, T. Blaine Crowley, Lena Dorfschmidt, Ayan S. Mandal, Dabriel Zimmerman, Remo M.S. Williams, Smrithi Prem, Elizabeth Levitis, Margaret Gardner, Katherine Cyr, Viveknarayanan Padmanabhan, Jerome H. Taylor, Kosha Ruparel, Rune Boen, Carrie E. Bearden, Christopher R. K. Ching, Bogdan Pasaniuc, Stewart Anderson, Daniel McGinn, Elaine Zackai, Beverly Emanuel, Sarah Hopkins, Madeline Chadehumbe, Karen J. Low, Tim J. Cole, Richard A.I. Bethlehem, R. Taki Shinohara, J. William Gaynor, David R. Roalf, Raquel E. Gur, Donna M McDonald-McGinn, Aaron Alexander-Bloch, ENIGMA 22q11.2 Deletion Syndrome Working Group

## Abstract

**Background:** 22q11.2 deletion syndrome (22q11DS) is a common microdeletion associated with widespread brain alterations and elevated risk for schizophrenia and other neuropsychiatric conditions. Prospective research studies often exclude individuals with severe cognitive impairment, medical comorbidities, or inability to tolerate research MRI without sedation, features common in 22q11DS. This limits both the generalizability of neuroimaging findings and our understanding of the full phenotypic spectrum. Moreover, while standard brain growth charts quantify deviation from typical development, they cannot identify patients who are disproportionately affected relative to their genetic peers, limiting clinical utility for risk stratification. Leveraging clinical MRI data offers a scalable approach to address these gaps.

**Methods:** We analyzed 92 patients with 22q11DS (age 0.5-21 years, 49% female) and 252 matched clinical controls. Using normative modeling derived from 1,995 reference clinical scans, we quantified individual-level brain deviations from population norms. We validated clinical findings against the independent ENIGMA-22q research consortium, characterized rates of extreme structural deviations to assess within-syndrome heterogeneity, correlated spatial patterns of brain alterations with gene expression from the Allen Human Brain Atlas, and generated syndrome-specific growth charts to test whether deviations from syndrome-specific norms predicted cognitive and language outcomes.

**Results:** Patients with 22q11DS showed widespread reductions in brain volumes (max Cohen’s d=−1.31) and cortical surface area (d=−0.71) with increased cortical thickness (d=0.39). These findings were highly convergent with the ENIGMA-22q research cohort (r=0.61-0.87). Forty percent of patients showed at least one global brain measure below the 2.5th percentile. Spatial patterns of cortical volume and surface area correlated with the expression of genes within the 22q11.2 locus. Critically, syndrome-specific growth charts revealed that smaller cerebellar volume relative to 22q11DS peers predicted lower language scores across two independent assessment methods (p<0.03), demonstrating potential prognostic utility.

**Conclusions:** This study provides a critical proof of principle for using heterogeneous clinical imaging to robustly characterize brain structure in rare genetic disorders. Syndrome-specific growth charts provide a novel framework to quantify within-syndrome variability and demonstrate potential prognostic value by linking individual brain structure to cognitive outcomes.

## Background

22q11.2 deletion syndrome (22q11DS; OMIM #188400, #192430) carries a profound burden of neuropsychiatric and cognitive symptoms [1]. Notably, schizophrenia occurs in approximately 25% of adults with 22q11DS, alongside anxiety, attention-deficit/hyperactivity disorder (ADHD), and autism spectrum disorders (ASD) [1]. The syndrome is also characterized by significant cognitive deficits spanning speech, language, executive function, and memory [2]. These neuropsychiatric symptoms are co-occurrent with a host of multi-system symptoms including thymic dysplasia leading to immunodeficiency, congenital heart defects, and hypocalcemia, highlighting the pleiotropic nature of the deletion [1].

The diverse symptoms of 22q11DS are accompanied by widespread differences in brain structure compared with typically developing controls. Neuroimaging studies have consistently identified widespread reductions in cortical and subcortical gray matter volumes (GMV), together with lower cortical surface area (SA) and higher cortical thickness (CT), in 22q11DS relative to controls [3–6]. Furthermore, inter-individual differences in brain structure have been linked to both cognitive ability and risk for psychosis [4,6–8]. Many of these structural differences were first robustly identified through prospective research studies and through mega-analyses conducted by the ENIGMA 22q11.2 Deletion Syndrome (ENIGMA-22q) working group [4,6,9].

Despite our current understanding of brain structure in 22q11DS, prospective research studies may not capture the full heterogeneity of the syndrome. Research studies frequently employ exclusion criteria that remove individuals with severe neurodevelopmental deficits, such as impaired language, lower IQ scores, or an inability to tolerate research MRI scans without sedation. Because these features are common in 22q11DS, their exclusion likely leads to a substantial loss of phenotypic heterogeneity and findings that may not generalize to the broader 22q11DS population [1]. For example, rates of incomplete hippocampal inversion are higher in clinical samples of 22q11DS than in research samples [10]. Furthermore, existing neuroimaging studies have not included infants and toddlers, a critical gap given that neuroanatomical differences in 22q11DS are thought to emerge early in life [4–6]. Repurposing clinical imaging for quantitative imaging studies may bridge this gap by capturing individuals often missed in traditional research, thereby revealing a broader spectrum of neuroanatomical phenotypes.

Characterizing neuroanatomical heterogeneity also requires moving beyond case-control comparisons. While case-control comparisons can reveal group differences, often obscured are the subset of patients who manifest extreme biological deviations [11]. Normative modeling enables an analytic shift through the creation of brain “growth charts” [12–14], which provide an age- and sex-adjusted measure of deviation from the population norm and allow for the identification of individuals with “extreme” deviations [15]. Quantifying these individual-level extremes is critical for understanding the heterogeneity within the syndrome, as extreme phenotypes may represent distinct etiological subgroups requiring tailored interventions. Research in first episode psychosis, for instance, demonstrates that cortical thickness deviations are associated with negative symptom severity [16].

However, standard growth charts only tell us that a patient with 22q11DS diverges from the reference population norm. To identify patients who are disproportionately affected relative to their genetic peers, we must establish syndrome-specific brain growth charts. Disorder-specific growth charts for head circumference in neurofibromatosis type 1 (NF1) have demonstrated the clinical utility of this approach, examining how lower head circumference relative to peers in NF1 relates to developmental delay [17]. By linking individual-level deviations to clinical outcomes, we provide a pathway towards identifying imaging markers that capture the syndrome’s profound clinical heterogeneity.

Furthermore, while the macroscopic neuroanatomy of 22q11DS is well-characterized, the cellular mechanisms driving these changes remain largely unknown. Imaging transcriptomics offers a unique opportunity to link these macroscopic brain changes to their molecular underpinning by testing if the specific genes within the 22q11.2 locus are spatially correlated with the observed structural deviations. Prior work in the ENIGMA-22q cohort has previously linked cortical alterations to expression of genes involved in corticogenesis and mitochondrial function [9], offering the opportunity to validate these molecular signatures in clinical populations and a substantially younger cohort.

Here we demonstrate the utility of reutilizing clinical neuroimaging in rare genetic disorder research. We assembled a cohort of 92 individuals with 22q11DS from the Children’s Hospital of Philadelphia (CHOP) and a large reference sample of clinical scans with limited imaging pathology (SLIP) [12,18]. First, we validate clinical MRI as a tool for studying 22q11DS by comparing findings between clinical and research cohorts. Second, we characterize the heterogeneity of 22q11DS by examining rates of extreme deviation from the norm. Third, we probe the molecular underpinnings of these structural alterations by testing whether brain deviation patterns spatially correspond to expression of genes within the 22q11.2 deletion locus. Finally, we develop syndrome-specific growth charts and test whether deviations relative to genetic peers predict clinical outcomes, a critical step toward establishing prognostic biomarkers.

## Methods and Materials

### Study Cohorts and Design

Prospective data including the CHOP/Penn sample of ENIGMA-22q were collected with informed consent under protocols reviewed by CHOP and Penn Institutional Review Boards (IRB 10-007848 and IRB 812481 respectively). Retrospective use of clinical brain MRIs was reviewed by CHOP IRB 20–017870 and determined to be exempt from further oversight because it consisted of secondary use of existing data with low risk to human subjects and because informed consent could not realistically be obtained without biasing the sample.

The patient cohort was selected from individuals with 22q11.2 deletions and brain MRIs collected from 2000–2020. To ensure genomic homogeneity and reduce potential confounding from atypical deletion sizes, we restricted the sample to the standard 3Mb deletion at positions LCR22A-D (A–D), which accounts for approximately 85% of all cases [1]. This is critical as atypical 22q11.2 deletions show distinct phenotypic profiles [6]. After quality control, the final sample consisted of 92 patients (**Supplemental Methods**, **Figure S1**). Clinical indications for scans included developmental delay, suspected seizure, and follow-up after cardiac surgery.

A reference group of 1,995 was assembled from a previously described cohort of CHOP patients whose brain MRIs were determined to have limited or no imaging pathology through interpretation of signed radiology reports [12,18]. 252 clinical controls were selected from this reference group for case-control analyses, matched to 22q11DS cases by sex, scanner, and closest age.

Primary findings were compared to the independent ENIGMA-22q dataset (242 individuals with A-D deletion, 277 typically developing controls) (see **Supplemental Methods**).

### Image processing

We used FreeSurfer’s recon-all-clinical pipeline (v7.4.1), which incorporates deep learning-based segmentation (SynthSeg+) to enable accurate brain parcellation across various scan resolutions, contrasts, and orientations [19–22]. From these surface reconstructions, we extracted estimates for global and regional measures of GMV, sGMV, SA, and CT, as well as global white matter volume (WMV), ventricular volume, and cerebellum volume. CT analyses were restricted to high-resolution magnetization-prepared rapid acquisition gradient-echo (MPRAGE) sequences in patients ≥2 years (N=40/92, 43%) due to unreliable CT extraction in younger children and lower-resolution sequences.

Severe brain pathologies are common in 22q11DS and may impact the ability to accurately segment the brain or introduce extreme brain features that may bias findings towards structural pathologies as opposed to continuous brain variation. As such, each scan session from 22q11DS patients was evaluated by a board-certified neuroradiologist (J.E.S.) with expertise in 22q11DS to exclude patients with pathology that could interfere with surface reconstruction. Automated quality control measures from SynthSeg+ and surface Euler numbers (a proxy for scan quality) were further used to exclude low-quality segmentations (see **Supplemental Methods**) [20,23].

Since clinical scans often include multiple yet variable imaging acquisitions, we adopted a "median-of-session" approach [20]. For each unique scanning session, we processed all available images. Rather than arbitrarily selecting one scan, we used all scans that passed quality control within a session and calculated the median value for each brain measure across scans. This approach maximizes the signal-to-noise ratio and sample size compared to the alternative approach of selecting a single representative imaging sequence (e.g. MPRAGE). For patients with multiple distinct scanning sessions (i.e., longitudinal data), we prioritized the earliest session that passed quality control for the primary cross-sectional analyses, but retained all longitudinal samples for growth chart construction (see **Syndrome-Specific Growth Charts**). ENIGMA-22q research scans were processed using recon-all in FreeSurfer (v5.3) for consistency with prior studies [4,6] (see **Supplemental Methods**).

### Normative Modeling

To quantify individual-level brain deviations, we constructed normative population models of brain development (“brain growth charts”) using a large reference sample of 1,995 clinical scan sessions with limited or no imaging pathology [12,18]. These SLIP growth charts were generated using generalized additive models for location, scale, and shape (GAMLSS) to model growth of each feature, with fixed effects to account for age and sex as well as a random intercept for scanner [24]. Resulting models were applied to 22q11DS patients to generate standardized deviation scores (analogous to z-scores for normally distributed data). The Lifespan Brain Chart Consortium (LBCC) growth charts were used to derive ENIGMA-22q deviation scores [13]. Normative modeling pipelines are discussed in detail in the **Supplement Methods**.

### Spatial Association with Gene Expression

To investigate potential molecular underpinnings of 22q11DS brain structure, we compared spatial patterns of regional deviation scores with brain-wide gene expression data from the Allen Human Brain Atlas (AHBA) [25]. The primary principal component of gene expression in the brain was downloaded and parcellated using the *neuromaps* package in python [26]. Vertex-based principal components were then aggregated using the Desikan-Killany atlas to generate gene expression maps at the same scale as brain deviation scores. Individual gene expression maps were generated and parcellated using the *abagen* package in python. Our primary analysis focused on the complete set of genes located within the 22q11.2 deletion boundaries (A-D region) available in the AHBA dataset [25,27], allowing us to test the specific hypothesis that the spatial topography of brain deviations tracks the expression gradients of the deleted genes (see **Supplemental Methods**).

### Syndrome-Specific Growth Charts

To model brain growth specifically *within* the 22q11DS population, we applied the LMSz method [15]. This approach uses standardized deviation scores (z-scores) from growth charts of typical development as input and refines distributional parameters to create syndrome-specific growth charts. This allows for the generation of robust syndrome-specific charts even with smaller sample sizes.

Deviation scores from individuals with 22q11DS were used to estimate linear equations for the mean and scale of the 22q11DS brain growth charts (see **Supplemental Methods**). In the subset of patients with longitudinal imaging (N = 19), we included all available scans, with observations weighted by the inverse of the number of scans per participant to account for repeated measures [28]. An iterative model selection procedure was used to exclude outliers (absolute residuals > 3.5) before reconstructing the final models. The final models yielded two outputs: (a) 22q11DS-specific deviation scores, quantifying how an individual deviates from their cohort norm, and (b) centile lines characterizing this distribution. By combining the SLIP and 22q11DS-adjusted models, we back-transformed the centile lines to generate population growth charts for 22q11DS. Growth charts were generated for global brain metrics and at the regional level using the Desikan-Killiany parcellation, while the primary exploratory analysis focused on global metrics.

### Cognitive and Language Data

A subset of electronic health records (EHR) in the 22q11DS cohort contained intelligence quotient (IQ) and language ability assessments. IQ and language scores in the EHR were matched to the closest clinical scan for evaluating inter-individual differences in brain structure and their relationship to cognitive outcomes.

IQ was assessed in 33 patients through the use of age-appropriate Weschler assessment scales, namely the Wechsler Preschool and Primary Scale of Intelligence [29,30], Wechsler Intelligence Scale for Children (WISC) [29,31,32], Wechsler Abbreviated Scale of Intelligence (WASI) [33], and the Wechsler Adult Intelligence Scale [34,35]. The Wechsler assessment scales evaluate intelligence quotients across multiple cognitive domains. Measures of IQ are further subdivided into subcategories of (VIQ) and (PIQ).

Language ability was evaluated through two independent scales: the Clinical Evaluation of Language Fundamentals (CELF; N = 48) [36,37] and the Preschool Language Scales (PLS; N = 58) [38–40]. The CELF is a structured assessment administered to individuals between the ages of six and 21 that provides standardized scores for core language, receptive language, and expressive language. The PLS is a play-based assessment designed for individuals up to seven years old and provides standardized scores for language, auditory comprehension, and expressive communication.

### Statistical Analysis

To assess case-control differences, each brain deviation score was predicted using a linear fixed-effects model. The primary predictor was diagnosis, with covariates for age, sex, and the median Euler number (a proxy for scan quality). Age and sex were included to account for any residual biases not fully captured by normative modeling. From these models, we converted t-statistics to Cohen’s d effect sizes [41] and corrected all p-values for multiple comparisons using the Benjamini-Hochberg FDR procedure [42].

We used a permutation test to assess the statistical significance of the spatial correlation (Pearson’s r) between the effect size maps from our clinical cohort and the ENIGMA-22q cohort. To do this, we first generated 1,000 null effect size maps using the Freedman-Lane procedure [43]. We then calculated correlations between these null effect size maps and the ENIGMA-22q map. The p-value was derived by comparing our observed correlation to this null distribution. As a confirmatory analysis, we also used a spatial "spin test" [44], which produced convergent results.

We defined an "extreme" measure as any value falling below the 2.5th or above the 97.5th percentile of the reference population. We then used a permutation test to determine if the rate of these extreme measures was significantly higher in patients with 22q11DS than would be expected by chance. This was done by creating a null distribution from 1,000 permutations of the case-control diagnosis labels and calculating the difference in extreme rates for each permutation.

To quantify the relationship between brain deviations and gene expression, we first calculated the Pearson correlation between the median brain deviation map and each gene’s expression map. We performed this analysis separately for patients with 22q11DS and for controls. We then used a permutation test to determine if the brain-gene correlation was significantly different between the two groups. A null distribution was generated by calculating the difference in correlation coefficients across 1,000 permutations of the diagnosis labels.

In an exploratory analysis, we used linear regression to investigate the relationship between an individual’s deviation from the 22q11DS-specific growth charts and their scores on IQ and language tests. Cortical thickness (CT) measures were excluded from these models due to the limited sample size for that measure.

## Results

### Cohort Characteristics

The primary clinical cohort consisted of 92 cases with 22q11DS (mean age=7.8 years; 49% female) and 252 matched clinical controls (**Table 1**). The ENIGMA-22q cohort (cases=242, controls=277) was significantly older (mean age=17.3 years) but did not differ significantly by sex or IQ (**Table S1**).

**Table 1.** Demographic characteristics.

|  | **22q11DS**<br>(N=92) | **CN**<br>(N=252) |  |
| --- | --- | --- | --- |
| **Age (years)** |  |  |  |
| Mean (SD) | 7.8 (5.2) | 8.8 (5.4) | t=-1.5, p=0.1 |
| Median [Range] | 7.3 [0.5, 20.3] | 7.6 [0.8, 19.6] |  |
| **Sex** |  |  |  |
| Female | 45 (49%) | 114 (45%) | $\chi^2=0.4$, p=0.5 |
| Male | 47 (51%) | 138 (55%) |  |
| **Race** |  |  |  |
| White | 67 (73%) | 143 (57%) | $\chi^2=8.1$, p=0.04 |
| Black/AA | 16 (17%) | 64 (25%) |  |
| Asian | 2 (2%) | 5 (2%) |  |
| Other/Multiple | 7 (8%) | 40 (16%) |  |
| **Ethnicity** |  |  |  |
| Non-Hispanic | 81 (88%) | 160 (63%) | $\chi^2=0.6$, p=0.4 |
| Hispanic | 11 (12%) | 29 (12%) |  |
| Unknown | 0 (0%) | 63 (25%) |  |
| **IQ** |  |  |  |
| Mean (SD) | 75.0 (13.0) | - | - |
| Median [Range] | 74 [52, 103] | - |  |
| Missing | 64 (70%) | - |  |

Data are presented as N (%) or mean (SD) and median [range]. P-values reflect the statistical comparison between the 22q11.2 deletion syndrome (22q11DS) patient group and the control group from the CHOP cohort. IQ scores were not available for the CHOP reference group. Abbreviations: CHOP, Children's Hospital of Philadelphia; CN, controls; IQ, intelligence quotient; SD, standard deviation.

### Global and Regional Deviations

Population modeling revealed widespread brain deviations in 22q11DS (**Figure 1**, **Table S1**). Patients with 22q11DS had smaller cerebellums (Cohen’s d=-1.31, p_fdr_<10^-25^), GMV (d=-0.72, p_fdr_<10^-9^), sGMV (d=-0.63, p_fdr_<10^-7^), WMV (d=-0.72, p_fdr_<10^-9^), and SA (d=−0.71, p_fdr_<10^-8^) compared to controls. Conversely, patients had larger ventricular volumes (d=0.52, p_fdr_<10^-6^) and higher mean CT (d=0.39, p_fdr_=0.02). Subcortical alterations were most prominent in ventral diencephalon and hippocampus (d=−0.75 to −0.83; p_fdr_<10^-9^) (**Figure 1B**). Cortical GMV and SA deviations were most pronounced in the caudal cortex (peak right cuneus d_GMV_=−1.32, d_SA_=−1.29; **Figure 1C**) with a gradient of attenuating effect sizes from caudal to rostral cortex. A prominent exception was higher insular volumes (d=0.28-0.39, p_fdr_<0.02). In contrast, CT deviations were largest in medial orbitofrontal cortex (d=0.89-0.99, p_fdr_<10^-7^) and did not demonstrate a gradient. Focal decreases were identified in left caudal anterior cingulate (d=-0.89, p_fdr_<10^-6^), parahippocampal (d=-0.62, p_fdr_<10^-3^), and superior temporal cortices (d=-0.38, p_fdr_=0.02). Clinical MRI effect sizes showed a significant correlation with ENIGMA-22q effect sizes (**Figure 1D**, **Figure S2**, **Table S2**) assessed using both permutation testing and a cortical “spin” test (r=0.61–0.87; p_perm_ < 0.002, p_spin_ < 0.001) [44]. While GMV and CT showed a strong 1-to-1 correspondence between cohorts, cortical SA effects were systematically more negative in ENIGMA-22q. Age- and sex-by-diagnosis interactions were largely non-significant (**Figures S3-S4**).

**Figure 1.**
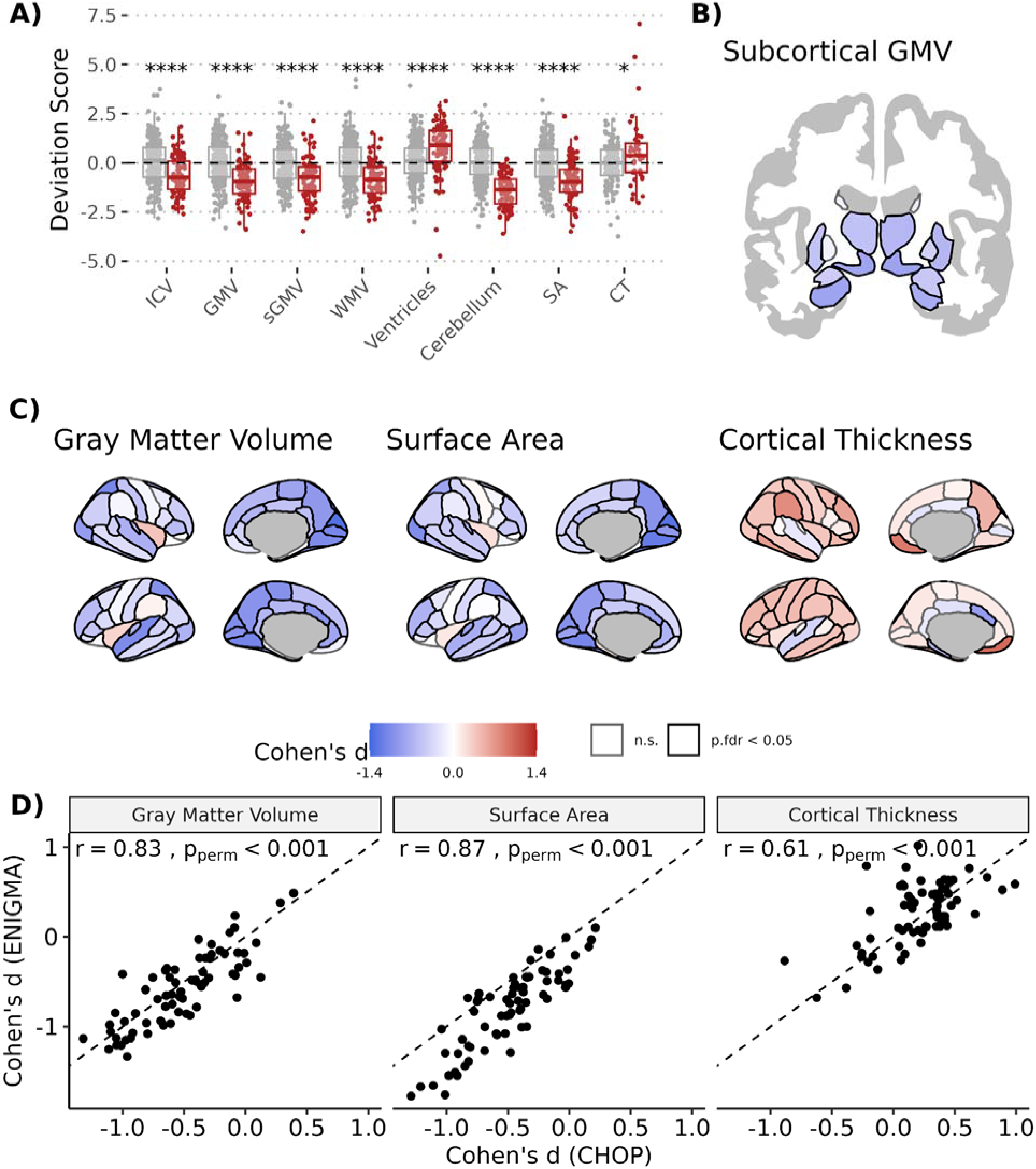
Effect sizes of 22q11.2 Deletion syndrome (22q11DS) on brain deviation scores. **(A)** Boxplots show individual standardized deviation scores for global brain measures in patients with 22q11DS (n=92) and controls (n=252). **(B** and **C)** Brain maps display Cohen’s d effect sizes for regional deviations in **(B)** subcortical gray matter volume (GMV) and **(C)** cortical GMV, surface area (SA), and thickness (CT). Regions with negative deviation scores in 22q11DS (i.e., smaller values in 22q11DS) are shown in blue, and regions with positive scores are shown in red. Solid black outlines indicate regions with significant case-control differences. **(D)** Scatter plots show the correlation between Cohen’s d effect sizes from the primary CHOP cohort (x-axis) and the independent ENIGMA-22q cohort (y-axis). All deviation scores were derived from population growth charts predicting deviations based on age, sex, and scanner. P-values were corrected for multiple comparisons using the Benjamini-Hochberg false discovery rate (FDR) procedure. Asterisks indicate the level of statistical significance after correction. **** p < 0.0001, *** p < 0.001, ** p < 0.01, * < 0.05. Abbreviations: CT, cortical thickness; GMV, gray matter volume; ICV, intracranial volume; n.s., not significant; SA, total SA; sGMV, subcortical GMV; WMV, white matter volume.

### Extreme Phenotypes

A substantial number of individuals with 22q11DS possessed brain features at the extremes of the reference distribution (**Table S4**). We found that 40% of cases had at least one global feature <2.5th percentile, while 21% had a global feature >97.5th percentile. The most frequent finding was low cerebellum volume, observed in 28% of 22q11DS. These extreme deviations extended to the regional level: a significant proportion of 22q11DS had features <2.5th percentile for the majority of GMV and SA, and >97.5th percentile for CT (**Figure 2**). The regional distribution of these extreme deviations was moderately correlated with the ENIGMA-22q cohort for GMV (r=0.66, p_perm_=0.005, p_spin_<0.001) and SA (r=0.81, p_perm_=0.001, p_spin_<0.001) (**Figures S5-6**, **Table S5**). Rates of extreme deviations were not significantly different between clinical and research cohorts.

**Figure 2.**
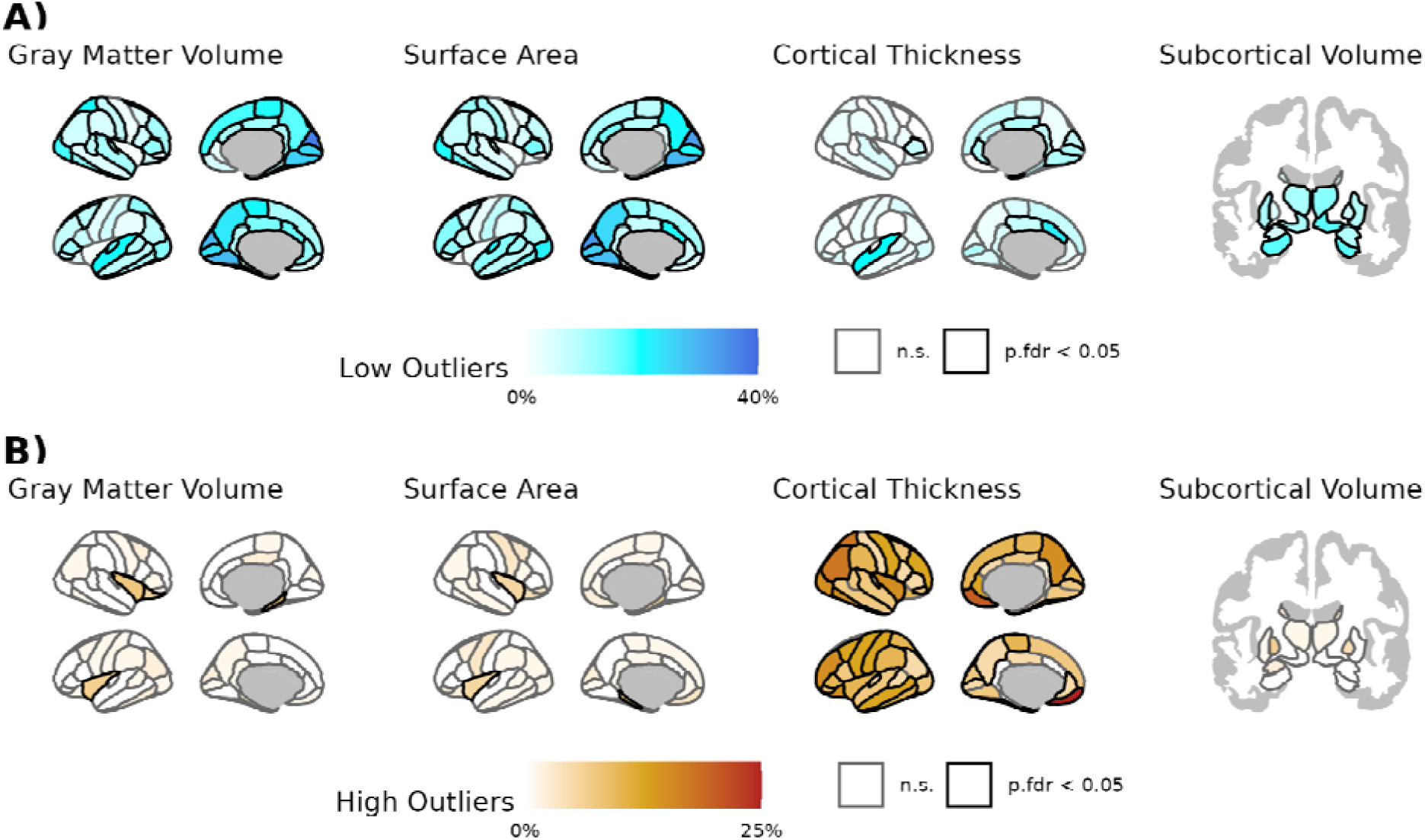
Extreme atypicality of brain features is common in 22q11.2 Deletion syndrome (22q11DS). Brain maps showing the percentage of patients with atypically low scores **(A)** and atypically high scores **(B)** for each brain feature. Atypically low scores were defined as falling below the 2.5^th^ percentile of the reference distribution, while atypically high scores were defined as above the 97.5^th^ percentile. The statistical significance of regional enrichment for extreme phenotypes (indicated by solid black outlines) was assessed using a permutation test of case-control diagnoses (1,000 permutations). P-values were corrected for multiple comparisons using the Benjamini-Hochberg false discovery rate (FDR) procedure. n.s. indicates not significant.

### Brain Deviation Patterns Correlate with Gene Expression

The median deviation maps of GMV and SA, but not CT, demonstrated significantly higher correlations with the first principal component of gene expression in 22q11DS compared to controls (**Figure 3A**; GMV: r_22q_=-0.50, r_control_=0.15, p_perm_=0.002; SA: r_22q_=-0.48, r_control_=0.19, p_perm_ =0.03). We identified six genes within the 22q11.2 locus with expression correlated to median deviation scores (**Figure 3B**, **Table S6**): *TXNRD2*, *P2RX6*, *C22orf39*, and *DGCR8* with GMV and *TXNRD2*, *AIFM3,* and *ZNF74* with SA. All gene correlations with SA and the *TXNRD2* correlation with GMV were also found in the ENIGMA-22q cohort (**Figure S7** and **Table S7**).

**Figure 3.**
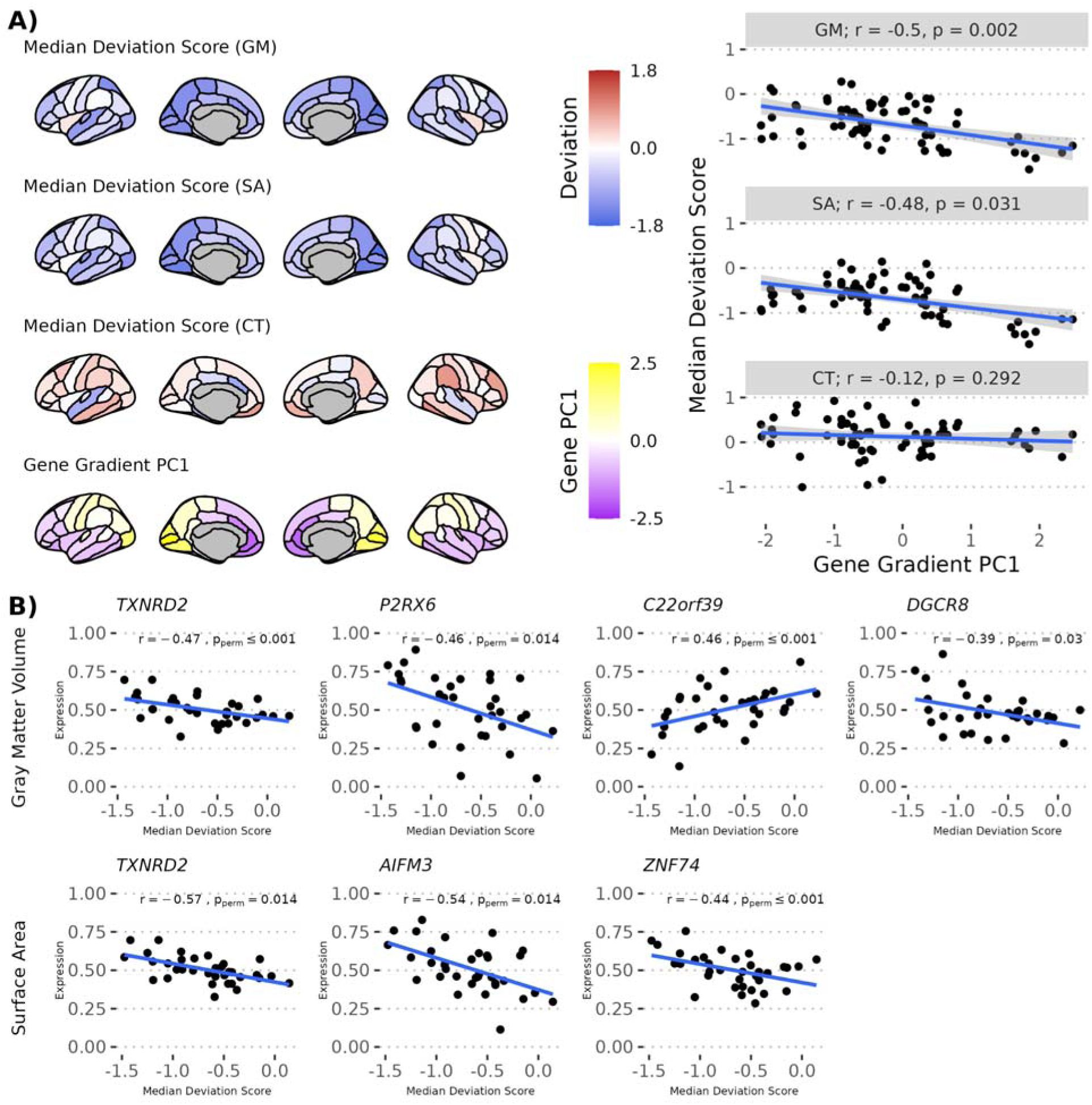
Spatial correlation between brain deviations in 22q11.2 Deletion syndrome (22q11DS) and expression of genes in the 22q11.2 locus in postmortem brain tissue. **(A)** The spatial maps of median deviation scores for gray matter volume (GMV), surface area (SA), and cortical thickness (CT) were compared to the first principal component (PC1) of brain-wide gene expression from the Allen Human Brain Atlas. The scatter plots on the right show a significant negative correlation between the PC1 gradient and the deviation maps for both GMV (r = −0.50, p_perm_ < 0.001) and SA (r = −0.48, p_perm_ = 0.03) in 22q11DS. No significant correlation was observed for CT (r = −0.12, p_perm_ = 0.3). **(B)** Follow-up analyses show significant correlations between the GMV and SA deviation maps and the expression patterns of individual genes in the 22q11DS locus. Shown are the six 22q11DS genes that were significantly correlated with deviation maps after multiple comparisons correction. Significance was assessed by comparing the difference in correlation coefficients of cases and controls to a null distribution across 1,000 permutations. Abbreviations: CT, cortical thickness; GMV, gray matter volume; PC1, first principal component; SA, surface area.

### Characterizing 22q11DS-specific Brain Growth Charts

Syndrome-specific 22q11DS brain growth charts revealed developmental trends that were shifted compared to typical development (**Figure 4A**, **Table S8**). At the global level, models showed lower medians for GMV, sGMV, WMV, cerebellum volume and SA as well as higher medians for ventricular volume, but these shifts were stable across age and sex. At the regional level, only subtle age and sex variations were observed (**Figure S8**).

**Figure 4.**
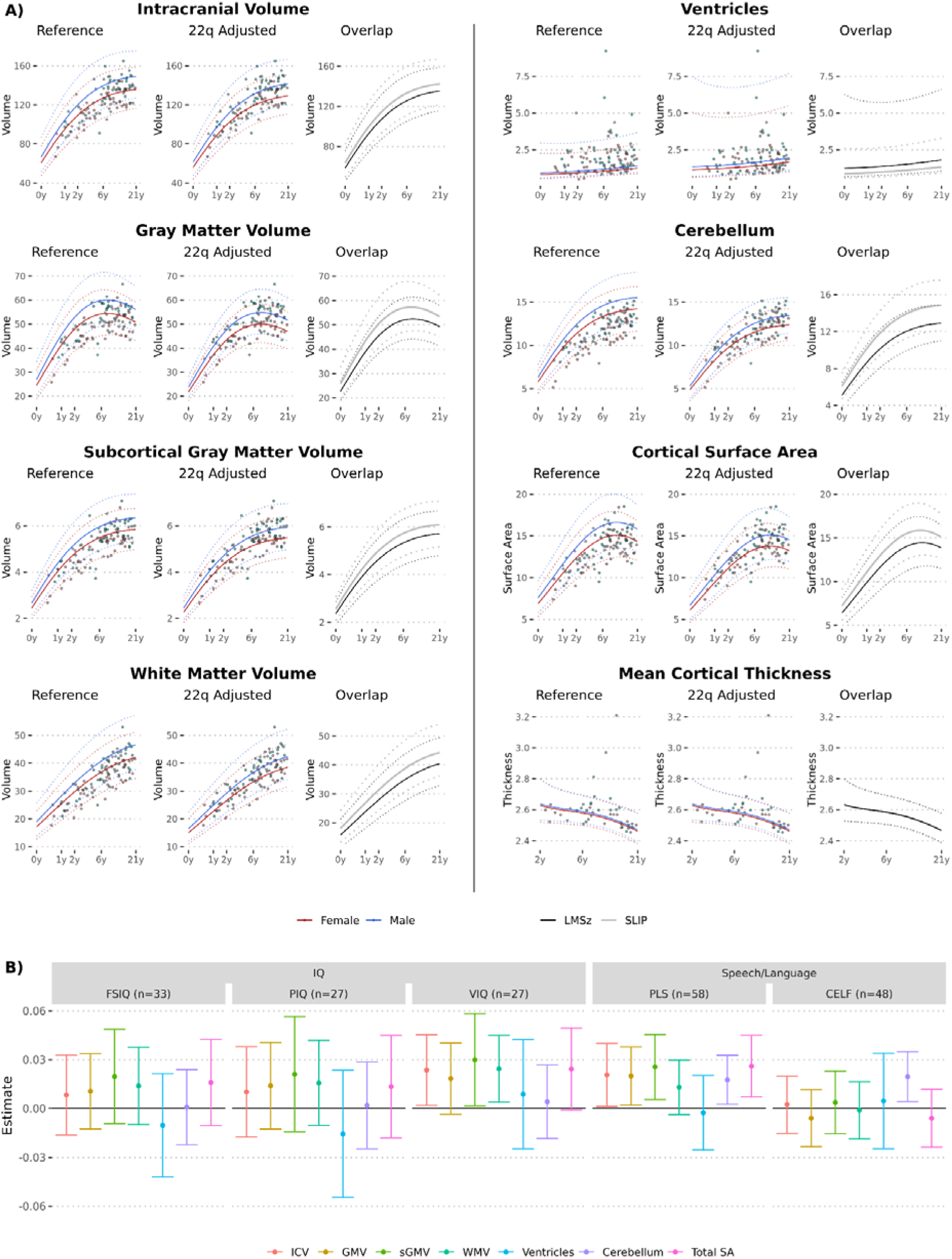
Syndrome-specific brain growth charts for 22q11.2 Deletion syndrome (22q11DS) and their association with cognitive outcomes. (A) Global brain growth charts for intracranial volume (ICV), cortical gray matter volume (GMV), subcortical GMV (sGMV) and white matter volume (WMV). For each measure, the left panel ("Reference") shows individual data points from the 22q11DS cohort plotted against the reference population’s growth curves. The middle panel ("22q Adjusted") displays the same data points against the newly generated growth charts that are specific to 22q11DS. The right panel ("Overlap") directly compares the median (solid lines) and 2.5th/97.5th centile (dashed lines) curves for both the reference (gray) and 22q11DS-specific models (black) averaged across sexes to visualize the shift in curves. (B) Exploratory analysis linking brain deviations to clinical outcomes. The points represent the estimated association (standardized beta coefficient) from linear models testing the relationship between an individual’s deviation from 22q11DS-specific growth charts and their scores on intelligence and language tests. Error bars indicate 95% confidence intervals. Cortical thickness measures were excluded from this analysis due to limited sample sizes. Abbreviations: CELF, Clinical Evaluation of Language Fundamentals; CT, cortical thickness; FSIQ, full-scale intelligence quotient; GMV, gray matter volume; ICV, intracranial volume; PIQ, performance intelligence quotient; PLS, Preschool Language Scales; SA, surface area; sGMV, subcortical GMV; VIQ, verbal intelligence quotient; WMV, white matter volume.

### Association of Brain Deviations with Cognitive and Language Outcomes

In an exploratory analysis, an individual’s deviation from 22q11DS brain charts was associated with verbal intelligence and language measures (**Figure 4B**). Higher verbal IQ (n=27) was nominally associated with intracranial volume (t=2.2, df=25, p=0.03), sGMV (t=2.2, df=25, p=0.04), and WMV (t=2.4, df=25, p=0.03). Higher proficiencies in both language assessments were associated with larger cerebellar volume (t=2.3, df=56, p=0.02 and t=2.6, df=46, p=0.01). Higher PLS scores were additionally associated with larger values across all global measures except ventricles (t=2.1-2.7, df=56, p<0.04).

### Sensitivity Analyses

We performed several sensitivity analyses, which confirmed the robustness of reported findings (see **Supplemental Methods**). Sensitivity analyses included: controlling for intracranial volume; using only high-resolution MPRAGE scans; using alternative brain measures (i.e., centile scores and “raw” brain features non-benchmarked to brain growth charts); including patients with clinically significant pathology on neuroradiological review; and limiting the clinical cohort to scans in patients 6 years or older. Findings were robust to all sensitivity analyses.

## Discussion

We demonstrate that heterogeneous clinical MRI data provide a powerful, scalable approach for studying brain structure in rare genetic disorders. Using robust analysis pipelines, we charted brain deviations in 22q11DS from the first year of life, characterized by reduced SA and GMV alongside increased CT. Our principal findings are fourfold: (1) the convergence of findings between clinical MRI and prospective research cohorts supports generalizability of research findings to a broader, more clinically representative 22q11DS population; (2) normative modeling highlights both the magnitude and the heterogeneity of deviations from typical brain development quantified by reference brain charts; (3) brain deviations in 22q11DS mirror expression patterns in postmortem brain tissue of genes within the 22q11.2 locus; and (4) individualized deviation scores relative to peers benchmarked to 22q11DS-specific brain growth charts may be associated with cognitive and language outcomes.

Normative modeling reveals both the magnitude and the heterogeneity of deviations from reference brain charts. Forty percent of the 22q11DS cohort possessed at least one global brain measure in the bottom 2.5th percentile, highlighting the severe and pervasive neurobiological impact of the deletion. This level of extreme atypicality contrasts sharply with studies of idiopathic schizophrenia or ASD, which report overlapping extreme deviations in rarely more than 10% of individuals [45–47]. However, considerable heterogeneity existed even within this genetically homogeneous cohort. No brain region showed extreme deviations in a majority of patients, indicating that other factors—including epigenetics, environmental effects, and background polygenic variation—are likely significant contributors to brain structure variability in 22q11DS. This profound heterogeneity demonstrates why group-level case-control comparisons are insufficient for clinical care; understanding an individual patient’s risk requires a personalized framework capable of benchmarking this variability. As brain structural heterogeneity may be associated with clinical outcomes (including language ability in this study and psychosis later in life [6]), continuing to unravel the multifactorial nature of brain heterogeneity in 22q11DS remains an important direction towards implementing clinical prediction and targeted interventions.

We did not observe higher rates of extreme deviations in the clinical cohort compared to ENIGMA-22q, which suggests that the brain phenotypes of clinical 22q11DS may not be significantly more severe than those of research samples. Two factors may explain this discrepancy. First, many research cohorts are recruited directly from specialty clinics, meaning both groups represent individuals seeking diagnosis and treatment, potentially narrowing the phenotypic gap. Second, our necessary exclusion of gross pathology (e.g., arachnoid cysts, tumors) to ensure accurate surface reconstruction may have removed the most extreme neuroanatomical phenotypes from the clinical sample. A future approach that retains individuals with pathology but masks the affected regions could investigate this possibility further.

Despite the well-characterized neuroanatomical signature of 22q11DS, the cellular underpinnings of macroscopic brain alterations remain unknown. Reduced cortical SA may reflect impaired proliferation of neuronal progenitor cells during gestation, thereby limiting the overall size of the cortical sheet [6,48]. Increased CT may reflect over-proliferation of intermediate progenitor cells, atypical neuronal migration, or abnormal cortical folding [48,49]. This hypothesis aligns with known early developmental origins in 22q11DS; the deletion causes dysmorphogenesis of the pharyngeal arch system during embryonic development, driving cardiac and craniofacial defects [1].

Hypothesized early developmental origins are also supported by our results, which suggest that deviations from typical brain development occur early in life and thereafter remain relatively stable. We observed no significant age-by-diagnosis interactions, and 22q11DS growth charts were largely stable across age, even with the inclusion of children as young as six months. This provides cross-sectional evidence that the primary neuroanatomical signatures of 22q11DS are established early and proceed along a relatively parallel, albeit offset, trajectory. This aligns with previous SA studies, although some studies found age-dependent effects on CT which we did not detect, likely reflecting reduced statistical power to detect subtle age-related slopes in a cross-sectional sample (n=40) compared to larger longitudinal cohorts [49,50].

The spatial correspondence between gene expression and brain deviations supports mitochondrial involvement. Our imaging-transcriptomic analysis implicates two mitochondrial genes: *TXNRD2* (essential for reactive oxygen species level regulation [51]) and *AIFM3* (involved in NADPH oxidation and mitochondrial apoptosis [52]). Diminished dosage of *Txnrd2* in mouse models mirrors the neuronal and mitochondrial deficits in layer 2/3 projection neurons of 22q11DS mouse models [51]. Furthermore, introduction of *Txnrd2* rescues the neuronal antioxidant and dendritic growth deficit in vitro. While mitochondrial dysfunction has been linked to cognitive issues and impaired neurogenesis, the relationship between mitochondrial dysfunction and cortical surface area is unclear [53]. Of note, genes related to mitochondrial function were previously implicated by an imaging-transcriptomics study conducted in ENIGMA-22q [9]. Importantly, these molecular findings suggest that mitochondrial dysfunction could be a unifying mechanism linking gene deletion to structural brain alterations and, potentially, to the clinical heterogeneity observed across patients.

A primary innovation is the development of syndrome-specific brain growth charts. While standard norms quantify deviation from typical biology, syndrome-specific norms benchmark severity within the disorder, supporting the move beyond group-level comparisons toward a more personalized clinical framework. Specifically, in exploratory analyses we identified a novel link between smaller cerebellar volume, relative to 22q11DS brain charts, and lower language proficiency scores across two different assessments. This finding connects cerebellar hypoplasia, a common feature of 22q11DS [3], to speech and language deficits characteristic of the syndrome. With further validation, clinicians could plot a child’s cerebellar volume on such charts. A value in a low percentile for the syndrome could inform placement into higher-intensity treatment groups, ensuring personalized treatment plans that optimize scarce clinical resources. Future development could additionally link early imaging biomarkers to long term outcomes in 22q11DS. To this end, combining early-life clinical imaging with prospective research assessing clinical, social, and cognitive outcomes may highlight prognostic biomarkers.

Critically, we highlight the wider potential for leveraging heterogeneous clinical MRI data to study rare genetic disorders. Given the large effect sizes typical of rare genetic disorders (e.g., cerebellar volume d=-1.3) and large availability of “clinical controls”, retrospective clinical cohorts achieve robust statistical power even with moderate sample sizes of genetic carriers. This approach helps overcome the recruitment barriers inherent to rare disorders; a recent systematic review of 58 CNVs associated with ASD or schizophrenia found that 66% lacked whole brain imaging studies with sufficient samples to adequately detect large effect sizes (z-score>1.2) [54]. The strong correlation with ENIGMA-22q validates this framework for scaling to additional pathogenic CNVs that have not received the same degree of research focus as 22q11DS.

Our study illustrates how clinical neuroimaging has the potential to advance rare disease research from being largely descriptive to yielding clinically actionable data. The convergence with research cohorts establishes that heterogeneous clinical data can robustly characterize neuroanatomy, while extreme deviation analysis quantifies the profound yet variable impact of the deletion. Molecular correlates provide mechanistic grounding, implicating mitochondrial dysfunction as a pathway linking genotype to phenotype. Syndrome-specific charts quantify this heterogeneity into clinically-actionable markers, and their association with language outcomes demonstrates proof-of-principle for prognostic utility. Together, this framework utilizes clinical imaging for validation, characterization, and clinical translation. By applying this scalable approach to the large number of other neuropsychiatric CNVs that currently lack prospective research cohorts [54], we can progress precision medicine across rare genetic disorders.

## Limitations

Several limitations warrant consideration. While our analytical approach was designed for image heterogeneity and converged with research data, subtle effects from clinical scan heterogeneity cannot be excluded. Additionally, segmentation was unreliable for infants under 6 months, likely due to early myelination-related contrast inversion [55], making this critical developmental period a target for technical development.

Brain structure in 22q11DS may be influenced by factors beyond the deletion itself. Our study utilized a retrospective clinical cohort, meaning that concurrent medication data and detailed comorbidity phenotypes were not uniformly available. Psychotropic medications, which are common in this population, have been associated with brain structural changes in other contexts [56], though their specific impact in 22q11DS remains an area of active investigation [4,6]. Furthermore, brain structure in 22q11DS may be mediated by co-occurrent symptoms or procedures. For example, corrective and palliative surgeries for congenital heart defects have been associated with impaired perioperative brain growth [57,58], although recent work using fetal MRI suggests that brain growth alterations in 22q11DS may precede corrective cardiac surgeries [59].

Additionally, because cognitive assessments were not collected concurrently with the MRI (often separated by months or years), the observed brain-behavior associations should be interpreted as reflecting a general developmental relationship rather than a state-dependent link. Finally, our analyses are cross-sectional and cannot make causal inferences about developmental trajectories, for which longitudinal data are required. Future prospective studies with tightly coupled imaging and phenotyping are needed to resolve these temporal dynamics.

## Conclusions

Notwithstanding these limitations, our study establishes a framework for neuroimaging research through 22q11DS that is scalable to additional genetic disorders. By combining large-scale clinical MRI data with advanced analytical techniques, we can overcome many traditional barriers to studying the brain in rare genetic conditions. Syndrome-specific growth charts may be a critical next step toward developing targeted, early interventions to improve outcomes for individuals affected by 22q11DS and other rare genetic disorders.

## Abbreviations

22q11DS: 22q11.2 deletion syndrome
ADHD: Attention-deficit/hyperactivity disorder
AHBA: Allen Human Brain Atlas
ASD: Autism spectrum disorders
CELF: Clinical Evaluation of Language Fundamentals
CHOP: Children’s Hospital of Philadelphia
CT: Cortical thickness
EHR: Electronic health records
ENIGMA-22q: ENIGMA 22q11.2 Deletion Syndrome working group
FDR: False discovery rate
FSIQ: Full-scale intelligence quotient
GAMLSS: Generalized additive models for location, scale, and shape
GMV: Gray matter volume
ICV: Intracranial volume
IQ: Intelligence quotient
LBCC: Lifespan Brain Chart Consortium
MPRAGE: Magnetization-prepared rapid acquisition gradient-echo
PC1: First principal component
PIQ: Performance intelligence quotient
PLS: Preschool Language Scales
SA: Surface area
sGMV: Subcortical gray matter volume
VIQ: Verbal intelligence quotient
WASI: Wechsler Abbreviated Scale of Intelligence
WISC: Wechsler Intelligence Scale for Children
WMV: White matter volume

## Declarations

### Consent for publication

Not applicable

### Availability of data and materials

The participant-level data cannot be made publicly available as they contain protected health information from pediatric patients. The code used for the project can be accessed at github.com/BGDlab.

### Authors’ contributions

**BJ**, **JES**, **JS**, **DMMM, REG, and AAB.** conceived and designed the study. **BJ** performed the analyses, prepared all figures, and wrote the original draft of the manuscript. **JES**, **JS, JMS**, **SK, LD**, **RTS, and RAIB.** contributed to the statistical methodology and code development.

**JES**, **TBC**, **ASM**, **DZ**, **RMSW**, **SP**, **EL**, **MG**, **KC**, **VP**, **JHT.**, **KR**, **RB**, **CEB**, **CRKC**, **BP**, **SA**, **DM**, **EZ**, **BE**, **SH**, **MC**, **KJL**, **TJC**, **DRR**, **JWG**, **REG**, and **DMMM** contributed to data acquisition, phenotypic characterization, and processing.

**REG**, **DMMM**, and **AAB** provided supervision and funding acquisition. All authors contributed to the interpretation of the data and critical revision of the manuscript for intellectual content. All authors read and approved the final manuscript.

## Acknowledgements

Members of the ENIGMA 22q11.2 Deletion Syndrome Working Group (excluding individually named authors of this work):

Jennifer K. Forsyth, Maria Jalbrzikowski, Leila Kushan, Hoki Fung, Charles Schleifer, Paul M. Thompson, Julio E. Villalon-Reina, Therese van Amelsvoort, Geor Bakker, Wendy R. Kates, Kevin M. Antshel, Wanda Fremont, Linda E. Campbell, Kathryn L. McCabe, Eileen Daly, Maria Gudbrandsen, Declan G. Murphy, Michael C. Craig, Kieran C. Murphy, Jacob Vorstman, Ania Fiksinski, Sanne Koops, Tony. J Simon, Naomi J. Goodrich-Hunsaker, Courtney A. Durdle, Anne S. Bassett, Eva W. C. Chow, Nancy J. Butcher, Joanne Doherty, Adam Cunningham, Marianne van den Bree, David E. J. Linden, Michael J. Owen, Hayley Moss, Samuel Chawner, Nicolas A. Crossley, Gabriela M. Repetto, Kathleen Angkustsiri, Fabio di Fabio, Marianna Frascarelli, Joachim Hallmayer, Allan L. Reiss, Ruth O’Hara

## References

1. McDonald-McGinn DM, Sullivan KE, Marino B, Philip N, Swillen A, Vorstman JAS, et al. 22q11.2 deletion syndrome. Nat Rev Dis Primers. 2015;1:15071. 10.1038/nrdp.2015.71

2. Gur RC, Bearden CE, Jacquemont S, Swillen A, van Amelsvoort T, van den Bree M, et al. Neurocognitive profiles of 22q11.2 and 16p11.2 deletions and duplications. Mol Psychiatry. Nature Publishing Group; 2025;30:379–87. 10.1038/s41380-024-02661-y

3. Schmitt JE, DeBevits JJ, Roalf DR, Ruparel K, Gallagher RS, Gur RC, et al. A Comprehensive Analysis of Cerebellar Volumes in the 22Q11.2 Deletion Syndrome. Biol Psychiatry Cogn Neurosci Neuroimaging. 2023;8:79–90. 10.1016/j.bpsc.2021.11.008

4. Ching CRK, Gutman BA, Sun D, Villalon Reina J, Ragothaman A, Isaev D, et al. Mapping Subcortical Brain Alterations in 22q11.2 Deletion Syndrome: Effects of Deletion Size and Convergence With Idiopathic Neuropsychiatric Illness. Am J Psychiatry. 2020;177:589–600. 10.1176/appi.ajp.2019.19060583

5. Schmitt JE, Vandekar S, Yi J, Calkins ME, Ruparel K, Roalf DR, et al. Aberrant Cortical Morphometry in the 22q11.2 Deletion Syndrome. Biol Psychiatry. 2015;78:135–43. 10.1016/j.biopsych.2014.10.025

6. Sun D, Ching CRK, Lin A, Forsyth JK, Kushan L, Vajdi A, et al. Large-scale mapping of cortical alterations in 22q11.2 deletion syndrome: Convergence with idiopathic psychosis and effects of deletion size. Mol Psychiatry. 2020;25:1822–34. 10.1038/s41380-018-0078-5

7. Gudbrandsen M, Daly E, Murphy CM, Blackmore CE, Rogdaki M, Mann C, et al. Brain morphometry in 22q11.2 deletion syndrome: an exploration of differences in cortical thickness, surface area, and their contribution to cortical volume. Sci Rep. Nature Publishing Group; 2020;10:18845. 10.1038/s41598-020-75811-1

8. Schmitt JE, Yi JJ, Roalf DR, Loevner LA, Ruparel K, Whinna D, et al. Incidental radiologic findings in the 22q11.2 deletion syndrome. AJNR Am J Neuroradiol. 2014;35:2186–91. 10.3174/ajnr.A4003

9. Forsyth JK, Mennigen E, Lin A, Sun D, Vajdi A, Kushan-Wells L, et al. Prioritizing Genetic Contributors to Cortical Alterations in 22q11.2 Deletion Syndrome Using Imaging Transcriptomics. Cereb Cortex. 2021;31:3285–98. 10.1093/cercor/bhab008

10. Roalf D, Atkins A, Czernuszenko A, Pecsok MK, McDonald-McGinn DM, Schmitt JE, et al. Presence, severity, and functional associations of incomplete hippocampal inversion in 22q11.2 deletion syndrome. Biological Psychiatry: Cognitive Neuroscience and Neuroimaging [Internet]. 2025 [cited 2025 July 7]; 10.1016/j.bpsc.2025.04.009

11. Marquand AF, Rezek I, Buitelaar J, Beckmann CF. Understanding Heterogeneity in Clinical Cohorts Using Normative Models: Beyond Case-Control Studies. Biological Psychiatry. 2016;80:552–61. 10.1016/j.biopsych.2015.12.023

12. Schabdach JM, Schmitt JE, Sotardi S, Vossough A, Andronikou S, Roberts TP, et al. Brain Growth Charts for Quantitative Analysis of Pediatric Clinical Brain MRI Scans with Limited Imaging Pathology. Radiology. 2023;309:e230096. 10.1148/radiol.230096

13. Bethlehem RAI, Seidlitz J, White SR, Vogel JW, Anderson KM, Adamson C, et al. Brain charts for the human lifespan. Nature. 2022;604:525–33. 10.1038/s41586-022-04554-y

14. Dorfschmidt L, White S, Gardner M, Bedford S, Ball G, Edwards AD, et al. Charting structural brain asymmetry across the human lifespan [Internet]. bioRxiv; 2025 [cited 2025 July 30]. p. 2025.07.21.665924. 10.1101/2025.07.21.665924

15. Low KJ, Foreman J, Hobson RJ, Kwuo H, Martinez-Cayuelas E, Almoguera B, et al. The LMSz method - an automatable scalable approach to constructing gene-specific growth charts in rare disorders. medRxiv. 2024;2024.08.19.24312213. 10.1101/2024.08.19.24312213

16. Worker A, Berthert P, Lawrence AJ, Kia SM, Arango C, Dinga R, et al. Extreme deviations from the normative model reveal cortical heterogeneity and associations with negative symptom severity in first-episode psychosis from the OPTiMiSE and GAP studies. Transl Psychiatry. 2023;13:373. 10.1038/s41398-023-02661-6

17. Mandal AS, Shinohara RT, Jung B, Gardner M, Akouri HE, Yerys BE, et al. NF1-Specific Growth Charts for Head Circumference Over the First 3 Years of Life. Neurology. 2026;106:e214480. 10.1212/WNL.0000000000214480

18. Zimmerman D, Mandal AS, Jung B, Buczek MJ, Schabdach JM, Karandikar S, et al. A systematic protocol to identify “clinical controls” for pediatric neuroimaging research from clinically acquired brain MRIs. bioRxiv. 2025;2025.06.25.661530. 10.1101/2025.06.25.661530

19. Iglesias JE, Billot B, Balbastre Y, Magdamo C, Arnold SE, Das S, et al. SynthSR: A public AI tool to turn heterogeneous clinical brain scans into high-resolution T1-weighted images for 3D morphometry. Sci Adv. 2023;9:eadd3607. 10.1126/sciadv.add3607

20. Billot B, Magdamo C, Cheng Y, Arnold SE, Das S, Iglesias JE. Robust machine learning segmentation for large-scale analysis of heterogeneous clinical brain MRI datasets. Proc Natl Acad Sci U S A. 2023;120:e2216399120. 10.1073/pnas.2216399120

21. Billot B, Greve DN, Puonti O, Thielscher A, Van Leemput K, Fischl B, et al. SynthSeg: Segmentation of brain MRI scans of any contrast and resolution without retraining. Med Image Anal. 2023;86:102789. 10.1016/j.media.2023.102789

22. Gopinath K, Greve DN, Das S, Arnold S, Magdamo C, Iglesias JE. Cortical analysis of heterogeneous clinical brain MRI scans for large-scale neuroimaging studies [Internet]. arXiv; 2023 [cited 2026 Jan 16]. 10.48550/arXiv.2305.01827

23. Rosen AFG, Roalf DR, Ruparel K, Blake J, Seelaus K, Villa LP, et al. Quantitative assessment of structural image quality. Neuroimage. 2018;169:407–18. 10.1016/j.neuroimage.2017.12.059

24. Rigby RA, Stasinopoulos MD, Heller GZ, De Bastiani F. Distributions for Modeling Location, Scale, and Shape: Using GAMLSS in R. 1st ed. Chapman and Hall/CRC; 2019.

25. Hawrylycz MJ, Lein ES, Guillozet-Bongaarts AL, Shen EH, Ng L, Miller JA, et al. An anatomically comprehensive atlas of the adult human brain transcriptome. Nature. 2012;489:391–9. 10.1038/nature11405

26. Markello RD, Hansen JY, Liu ZQ, Bazinet V, Shafiei G, Suarez LE, et al. neuromaps: structural and functional interpretation of brain maps. Nat Methods. 2022;19:1472–9. 10.1038/s41592-022-01625-w

27. Markello RD, Arnatkeviciute A, Poline J-B, Fulcher BD, Fornito A, Misic B. Standardizing workflows in imaging transcriptomics with the abagen toolbox. eLife. eLife Sciences Publications, Ltd; 2021;10:e72129. 10.7554/eLife.72129

28. World Health Organization. WHO child growth standards: length/height-for-age, weight-for-age, weight-for-length, weight-for-height and body mass index-for-age: methods and development [Internet]. [cited 2025 Aug 25]. https://www.who.int/publications/i/item/924154693X. Accessed 25 Aug 2025

29. Wahlstrom D, Raiford SE, Breaux KC, Zhu J, Weiss LG. The Wechsler Preschool and Primary Scale of Intelligence—Fourth Edition, Wechsler Intelligence Scale for Children— Fifth Edition, and Wechsler Individual Achievement Test—Third Edition. Contemporary intellectual assessment: Theories, tests, and issues, 4th ed. New York, NY, US: The Guilford Press; 2018. p. 245–82.

30. Wechsler D. Wechsler Preschool and Primary Scale of Intelligence--Third Edition [Internet]. American Psychological Association (APA); 2012 [cited 2025 July 15]. 10.1037/t15177-000

31. Wechsler D. Wechsler Intelligence Scale for Children. 5th ed. Bloomington, MN: Pearson; 2014.

32. Wechsler D. Wechsler Intelligence Scale for Children, Fourth Edition [Internet]. American Psychological Association (APA); 2012 [cited 2025 July 15]. 10.1037/t15174-000

33. Wechsler D. WASI-II: Wechsler abbreviated scale of intelligence. PsychCorp; 2011.

34. Wechsler D. Wechsler Adult Intelligence Scale--Third Edition [Internet]. American Psychological Association (APA); 2019 [cited 2025 July 15]. 10.1037/t49755-000

35. Wechsler D. Wechsler Adult Intelligence Scale--Fourth Edition [Internet]. American Psychological Association (APA); 2012 [cited 2025 July 15]. 10.1037/t15169-000

36. Semel E, Wiig E, Secord W. Clinical Evaluation of Language Fundamentals, Fourth Edition. 2003.

37. Semel E, Wiig E, Secord W. Clinical Evaluation of Language Fundamentals, Fifth Edition. 2013.

38. Zimmerman IL, Steiner VG, Pond RE. Preschool Language Scale, Third Edition. San Antonio, TX: Psychological Corporation; 1992.

39. Zimmerman IL, Steiner VG, Pond RE. Preschool Language Scale, Fifth Edition. San Antonio, TX: Psychological Corporation; 2011.

40. Zimmerman IL, Steiner VG, Pond RE. Preschool Language Scale, Fourth Edition. San Antonio, TX: Psychological Corporation; 2002.

41. Ben-Shachar MS, Makowski D, Lüdecke D, Patil I, Wiernik BM, Thériault R, et al. effectsize: Indices of Effect Size [Internet]. 2025 [cited 2025 July 16]. https://cran.r-project.org/web/packages/effectsize/index.html. Accessed 16 July 2025

42. Benjamini Y, Hochberg Y. Controlling the False Discovery Rate: A Practical and Powerful Approach to Multiple Testing. Journal of the Royal Statistical Society: Series B (Methodological). 1995;57:289–300. 10.1111/j.2517-6161.1995.tb02031.x

43. Freedman D, Lane D. A Nonstochastic Interpretation of Reported Significance Levels. Journal of Business & Economic Statistics. ASA Website; 1983;1:292–8. 10.1080/07350015.1983.10509354

44. Alexander-Bloch A, Shou H, Liu S, Satterthwaite TD, Glahn DC, Shinohara RT, et al. On testing for spatial correspondence between maps of human brain structure and function. Neuroimage. 2018;178:540–51. 10.1016/j.neuroimage.2018.05.070

45. Bedford SA, Lai MC, Lombardo MV, Chakrabarti B, Ruigrok A, Suckling J, et al. Brain-Charting Autism and Attention-Deficit/Hyperactivity Disorder Reveals Distinct and Overlapping Neurobiology. Biol Psychiatry. 2024; 10.1016/j.biopsych.2024.07.024

46. Haukvik UK, Wolfers T, Tesli N, Bell C, Hjell G, Fischer-Vieler T, et al. Individual-level deviations from normative brain morphology in violence, psychosis, and psychopathy. Transl Psychiatry. 2025;15:118. 10.1038/s41398-025-03343-1

47. Wolfers T, Doan NT, Kaufmann T, Alnaes D, Moberget T, Agartz I, et al. Mapping the Heterogeneous Phenotype of Schizophrenia and Bipolar Disorder Using Normative Models. JAMA Psychiatry. 2018;75:1146–55. 10.1001/jamapsychiatry.2018.2467

48. Rakic P. Evolution of the neocortex: Perspective from developmental biology. Nat Rev Neurosci. 2009;10:724–35. 10.1038/nrn2719

49. Bagautdinova J, Zöller D, Schaer M, Padula MC, Mancini V, Schneider M, et al. Altered cortical thickness development in 22q11.2 deletion syndrome and association with psychotic symptoms. Mol Psychiatry. 2021;26:7671–8. 10.1038/s41380-021-01209-8

50. Jalbrzikowski M, Lin A, Vajdi A, Grigoryan V, Kushan L, Ching CRK, et al. Longitudinal trajectories of cortical development in 22q11.2 copy number variants and typically developing controls. Mol Psychiatry. Nature Publishing Group; 2022;27:4181–90. 10.1038/s41380-022-01681-w

51. Fernandez A, Meechan DW, Karpinski BA, Paronett EM, Bryan CA, Rutz HL, et al. Mitochondrial Dysfunction Leads to Cortical Under-Connectivity and Cognitive Impairment. Neuron. 2019;102:1127–1142.e3. 10.1016/j.neuron.2019.04.013

52. Xie Q, Lin T, Zhang Y, Zheng J, Bonanno JA. Molecular cloning and characterization of a human AIF-like gene with ability to induce apoptosis. J Biol Chem. 2005;280:19673–81. 10.1074/jbc.M409517200

53. Khacho M, Clark A, Svoboda DS, MacLaurin JG, Lagace DC, Park DS, et al. Mitochondrial dysfunction underlies cognitive defects as a result of neural stem cell depletion and impaired neurogenesis. Hum Mol Genet. 2017;26:3327–41. 10.1093/hmg/ddx217

54. Modenato C, Martin-Brevet S, Moreau CA, Rodriguez-Herreros B, Kumar K, Draganski B, et al. Lessons Learned From Neuroimaging Studies of Copy Number Variants: A Systematic Review. Biological Psychiatry. 2021;90:596–610. 10.1016/j.biopsych.2021.05.028

55. Dietrich RB, Bradley WG, Zaragoza EJ, Otto RJ, Taira RK, Wilson GH, et al. MR evaluation of early myelination patterns in normal and developmentally delayed infants. AJR Am J Roentgenol. 1988;150:889–96. 10.2214/ajr.150.4.889

56. Voineskos AN, Mulsant BH, Dickie EW, Neufeld NH, Rothschild AJ, Whyte EM, et al. Effects of Antipsychotic Medication on Brain Structure in Patients With Major Depressive Disorder and Psychotic Features: Neuroimaging Findings in the Context of a Randomized Placebo-Controlled Clinical Trial. JAMA Psychiatry. 2020;77:674–83. 10.1001/jamapsychiatry.2020.0036

57. Matthiesen NB, Agergaard P, Henriksen TB, Bach CC, Gaynor JW, Hjortdal V, et al. Congenital Heart Defects and Measures of Fetal Growth in Newborns with Down Syndrome or 22q11.2 Deletion Syndrome. J Pediatr. 2016;175:116–122.e4. 10.1016/j.jpeds.2016.04.067

58. Cromb D, Bonthrone AF, Maggioni A, Cawley P, Dimitrova R, Kelly CJ, et al. Individual Assessment of Perioperative Brain Growth Trajectories in Infants With Congenital Heart Disease: Correlation With Clinical and Surgical Risk Factors. Journal of the American Heart Association. Wiley; 2023;12:e028565. 10.1161/JAHA.122.028565

59. Cromb D, Finck T, Bonthrone AF, Uus A, Van Poppel M, Steinweg J, et al. An exploratory fetal MRI study examining the impact of 22q11.2 microdeletion syndrome on early brain growth. J Neurodev Disord. 2025;17:7. 10.1186/s11689-025-09594-9

