## Supplement Files 1 for "Charting Brain Structure in 22q11.2 Deletion Syndrome with Clinical Neuroimaging"

Table of Contents

***Supplementary Methods	2***

**Dataset characteristics and processing	2**

**Generation of deviation scores	3**

**Imaging transcriptomics	4**

**Generation of 22q11DS-specific growth charts	5**

**Sensitivity analyses	5**

***Supplementary Figures	8***

***References	28***

### Supplementary Methods

#### Dataset characteristics and processing

##### Children’s Hospital of Philadelphia (CHOP) Cohort

The primary dataset for this analysis consists of electronic health records and imaging collected between 2000 and 2020 from the Children’s Hospital of Philadelphia (CHOP). Patients were selected for inclusion in this study via records provided by the 22q and You Center, the largest program in the USA dedicated to the diagnosis and treatment of children with 22q11DS, having evaluated over 1,500 patients for the syndrome (<https://www.chop.edu/centers-programs/22q-and-you-center>).

Relevant clinical images from 22q11DS patients were collected using Arcus resources hosted at CHOP [1]. Arcus is a suite of tools and services developed to enhance research efforts at CHOP to explore available data, see overlaps among datasets, build new cohorts, and determine if there are data or samples available for additional research projects. Arcus connects CHOP's clinical and research data to enable reproducible research within a managed, scalable framework. This framework includes: 1) user access controls; 2) patient privacy and confidentiality protections through regulatory review; 3) electronic honest-brokered data de-identification and re-identification; and 4) data retention, management, sharing, and destruction services in an audited computational environment. Using 22q11DS patient identifiers, an honest-broker interfaced with the CHOP Department of Radiology to transfer clinical scans from the CHOP Picture Archiving and Communications Systems (PACS) to an internal research PACS. A deidentification pipeline, Locutus, was then employed to deidentify and deliver DICOM files for downstream processing (<https://github.com/BGDlab/Locutus>) [2]. Heudiconv and CuBIDS were used to convert the DICOM files into a BIDS directory of NIFTI files (<https://github.com/BGDlab/chop-bgd-image-curation>) [3,4].

While the request for clinical imaging data was initially encompassing of any individual with 22q11DS, we refined our selection to only those with a confirmed A-D deletion, which constitute around 85% of 22q11DS cases and allowed for our investigation to focus on a more genetically homogeneous cohort [5,6]. We describe the inclusion and exclusion of patients in **Figure S1**. First, an initial request for clinical scans returned 262 patients across 373 scan sessions. Of these, scans from 229 patients were successfully converted to BIDS format and found to have structural MRI scans of the brain. A total of 1,450 non-contrast scans were converted across 333 scan sessions (**Table S9**). These scans were each segmented independently using the recon-all-clinical pipeline in FreeSurfer v7.4.1 [7–10]. Automated quality control (QC) procedures were used to further subset scans, leveraging Euler numbers from recon-all-clinical and QC scores from SynthSeg+ to exclude low-quality scans and segmentations, excluding any scan with a mean Euler number < -60 or any SynthSeg+ Dice score < 0.65 [8]. We identified 871 scans (60% of the initial scans) across 223 scan sessions of 160 patients that passed both SynthSeg QC and Euler number-based thresholding. In total, 18 patients were excluded for pathology that could impact the ability to accurately segment the brain or introduce extreme brain measures that could bias findings towards structural pathologies as opposed to continuous brain variation. Examples of pathologies that were excluded included polymicrogyria and ventriculomegaly (**Figure S9**).

Given CHOP’s specialization as a pediatric hospital, most patients were 21 years old or younger. We thus excluded three individuals that were significantly older than the remaining cohort (age > 21 years). For primary analyses, we prioritized the earliest scan session that passed quality control. Since clinical visits often include multiple imaging acquisitions, we calculated the median value for each brain metric across all scans within that session that passed automated quality control. This approach maximizes the signal-to-noise ratio for that specific timepoint.

The attrition from the initial clinical query (N=262) to the final analytical sample (N=92) was driven by three distinct factors: (1) Forty requested individuals had no usable structural imaging upon retrieval. This reflects the inherent imprecision of clinical data extraction and potential causes may include scans cancelled prior to appointment, aborted mid-session, or archiving errors. (2) As scans were not collected with research optimization in mind, sequence parameters varied significantly. Protocol heterogeneity was a critical driver of attrition; for example, standardized MPRAGE sequences were excluded at a rate of 33% whereas older or non-standard sequences failed at significantly higher rates (**Figure S10**). Scanner strength was a key determinant of data inclusion, with 3T scans being excluded at lower rates than 1.5T scans (33% vs 44%) (**Figure S11A**). (3) We used a board-certified neuroradiologist (J.E.S.) to exclude gross pathology that would invalidate surface reconstruction (e.g., large arachnoid cysts). Scans in patients with exclusionary pathology were excluded by automated QC at higher rates than those without (58% vs 33%) (**Figure S11B**). Importantly, sensitivity analyses demonstrated that results were highly similar when including scans with pathology that passed automated QC, suggesting that automated QC may be sufficient to exclude segmentation-altering pathology. Finally, the removal of individuals older than 21 years or with non-standard (non A-D) deletions represents a methodological choice to maximize genetic and phenotypic homogeneity rather than a data quality issue.

A reference group was assembled from the same hospital system, comprising individuals whose brain MRIs were determined to have no or limited imaging pathology, using a previously described protocol [11,12]. This determination was made by two experienced graders with high inter-rater reliability (>0.85) through the interpretation of signed radiology reports from the CHOP Department of Radiology [11,12]. Individuals with excessive motion or artifacts from dental hardware noted in the radiology report were excluded. For case-control analyses, two to three individuals from the reference group were matched to each 22q11DS patient by sex, scanner, and closest age.

When multiple scans passing quality control were present in a single scan session, the median value across all scans that passed quality control was used for analysis. Since 2008, CHOP has deployed a harmonized magnetization-prepared rapid acquisition gradient-echo (MPRAGE) sequence for routine brain MRI examinations. This sequence produces high-quality images and is common in research studies [13]. Thus, as a sensitivity analysis, we used a cohort of high-resolution (≤ 1.5 mm in all dimensions) MPRAGE scans (N = 48). Manual evaluation of segmentations revealed suboptimal CT extraction in non-MPRAGE and in scans from children younger than two years old. Consequently, all analyses of CT features were restricted to a subset of MPRAGE scans from participants older than two years (N = 40).

##### ENIGMA-22q Working Group Cohort

Primary findings were compared to an independent dataset from the ENIGMA-22q working group. These T1-weighted scans were segmented using FreeSurfer recon-all (version 5.3) as described in previously published manuscripts [14–16]. For an in-depth description of this cohort, we refer readers to the primary ENIGMA-22q publications [15,16]. After selecting scanners that provided data for both controls and individuals with a confirmed A-D deletion, we retained a final sample of 242 participants with 22q11DS and 277 controls across nine scanners (**Table S10**). For modeling purposes, each scanner was treated as an independent site. Of note, seven subjects were present in both the CHOP clinical cohort and the University of Pennsylvania’s contribution to the ENIGMA-22q cohort. As the scans were collected at different facilities under different protocols and the goal was to assess consistency between clinical and research data, these seven participants were retained in both cohorts.

##### Defining composite brain features

While most brain features were direct outputs of the processing pipelines, the following three global features were created as composites of several smaller regions:

- Subcortical GMV was defined as the sum of the bilateral thalamus, caudate, putamen, pallidum, hippocampus, amygdala, accumbens, and ventral diencephalon.
- Cerebellum Volume was defined as the sum of the GMV and white matter cerebellar volumes.
- Ventricle volume was defined as the sum of the left lateral ventricle, right lateral ventricle, left inferior lateral ventricle, right inferior lateral ventricle, 3rd ventricle, and 4th ventricle.

#### Generation of deviation scores

We leveraged two previously developed reference brain growth charts to calculate brain deviation scores: one built from clinical scans with limited imaging pathology (SLIP) for the CHOP cohort[12], and one from the Lifespan Brain Chart Consortium (LBCC) for the T1-weighted ENIGMA-22q research scans [17,18]. Both sets of charts were constructed using generative additive models for location, scale, and shape (GAMLSS), which generate nonlinear, cross-sectional curves of brain development [19]. As described previously, the generalized gamma distribution was selected for all models, allowing for the modeling of three distributional parameters, after distribution family selection based on minimizing Akaike Information Criterion (AIC) and Bayesian Information Criterion (BIC). While these parameters are not identical to the statistical moments in the generalized gamma distribution, we follow the GAMLSS convention of referring to them as mean (µ), scale (σ), and skewness (ν). To harmonize the heterogeneous, multi-site data, scanner-specific random effects were modeled for both *mu* and *sigma*. The final models also included fixed-effect covariates for sex and log-transformed post-conception age. The LBCC models, which were run independently on multiple processing pipelines additionally contained a fixed covariate for segmentation software version. Post-conception age was computed by adding the available gestational age to the patient's age at scan; a default of 280 days was used when gestational age was unavailable. Consistent with our previous growth charts, the models used fractional polynomials to model nonlinear associations with age, with second or third order polynomials. Model selection was performed by minimizing BIC [12,17,18]. To apply these existing models to the new scanners in this study, we estimated scanner-specific random effects using a conditional maximum likelihood estimator [17,18]. Random effects were estimated using control samples from each site before being applied to 22q11DS samples. This approach allows for the generation of centile scores for out-of-sample measurements, which were then converted into “standardized deviation scores” using the qnorm function in R (equivalent to z-scores for normally distributed data). The specific application of each model is described below.

##### SLIP Clinical Growth Charts

The SLIP growth charts were used to model normative brain growth in the heterogeneous clinical data from our CHOP cohort. We extended the original SLIP pipeline, which was built on T1-weighted MPRAGE scans, to accept multimodal clinical data processed with the recon-all-clinical pipeline in FreeSurfer v7.4.1 [7–10,12]. We implemented a two-step, automated QC procedure, excluding any scan with a mean Euler number < -60 or any SynthSeg+ Dice score < 0.65. Since many scan sessions contained multiple modalities, we derived a median feature value for each session across all scans that passed QC, rather than prioritizing a single scan, to maximize sample size and signal-to-noise, following a previously published protocol [8].

The SLIP growth charts were constructed using brain features from 1,995 clinical scan sessions across 1,885 patients determined to have limited imaging pathology (mean age = 12.5 years, SD = 5.4, range = 0.26–37.5 years; 54% female) (https://github.com/BGDlab/chop-bgd-image-curation). For sensitivity analyses, we additionally generated growth charts solely from high-resolution, T1-weighted MPRAGE scans (sessions = 1,268, patients = 1,222, mean age = 12.8 years, SD = 5.3, range = 0.5–33 years; 55% female). Finally, for cortical thickness, we further subsetted to include only individuals greater than two years old (sessions = 1,225, patients = 1,185, mean age = 13.2 years, SD = 4.9, range = 2–33 years; 55% female). Model parameters are described in **Table S11**.

##### Lifespan Brain Consortium (LBCC) Reference Growth Charts

The LBCC growth charts were used to model normative brain growth in the T1-weighted research scans from the ENIGMA-22q cohort. These charts were generated from over 100,000 scans across 100 studies [17]. The original LBCC model generated bilateral average growth charts from 100 studies and more than 100,000 scans [17]. A notable advance from the original LBCC model is the extension of the models to contain hemisphere-specific data, with further details on this procedure described in Dorfschmidt et al. (2025) [18]. Model selection used the same approach as for the SLIP clinical growth charts. Model parameters are described in **Table 12**.

#### Imaging transcriptomics

##### Gene expression parcellation and processing

Regional microarray expression data were obtained from 6 post-mortem brains (1 female, ages 24.0–57.0, 42.50 +/- 13.38) provided by the Allen Human Brain Atlas (AHBA, <https://human.brain-map.org>) [20]. Data were processed with the abagen toolbox (version 0.1.4+15.gdc4a007; https://github.com/rmarkello/abagen) using an 83-region volumetric atlas in MNI space.

First, microarray probes were reannotated using data provided by Arnatkevic̆iūtė et al. (2019)[21]; probes not matched to a valid Entrez ID were discarded. Next, probes were filtered based on their expression intensity relative to background noise[22], such that probes with intensity less than the background in >=50.00% of samples across donors were discarded. When multiple probes indexed the expression of the same gene, we selected and used the probe with the most consistent pattern of regional variation across donors (i.e., differential stability [23]).

Here, regions correspond to the structural designations provided in the ontology from the AHBA. The MNI coordinates of tissue samples were updated to those generated via non-linear registration using the Advanced Normalization Tools (ANTs; https://github.com/chrisfilo/alleninf). Samples were assigned to brain regions in the provided atlas if their MNI coordinates were within 2 mm of a given parcel. To reduce the potential for misassignment, sample-to-region matching was constrained by hemisphere and gross structural divisions (i.e., cortex, subcortex/brainstem, and cerebellum, such that, e.g., a sample in the left cortex could only be assigned to an atlas parcel in the left cortex) [21]. All tissue samples not assigned to a brain region in the provided atlas were discarded. Inter-subject variation was addressed by normalizing tissue sample expression values across genes using a robust sigmoid function [24].

Gene expression values were then normalized across tissue samples using an identical procedure. Samples assigned to the same brain region were averaged separately for each donor and then across donors. As a proxy for brain expression, only genes within the top 50% of differential stability across donors were used for analyses [25,26]. Additionally, only the left hemisphere (i.e., the hemisphere with the greater sample size; N = 6 vs N = 2) was used for analyses. Of the 7,817 brain-expressed genes, we observed 1,269 genes whose expression patterns were significantly associated with the GMV map and 787 genes for the SA map after correction for multiple comparisons. Our analysis was restricted to the genes within the 22q11.2 deletion region (LCR A-D) that were adequately captured by the AHBA microarray probes.

#### Generation of 22q11DS-specific growth charts

We applied the LMSz method for modeling growth in rare genetic disorders leveraging reference growth charts [27]. We first generated a series of linear equations for mean (µ) and scale (σ) to characterize the relationship between age, sex, and deviation scores in 22q11DS (**Equations S1 and S2**) using the Normal (NO) family in GAMLSS [19]. The LMSz approach then fits all models and selects the model with the lowest BIC to generate adjusted growth charts.

$$deviation \sim age +sex+age*sex+1$$

$$deviation \sim age +sex+1$$

$$deviation \sim age+1$$

$$deviation \sim sex+1$$

$$deviation \sim1$$

$$deviation \sim0$$

**Equation S1.** Models for the µ parameter in 22q11DS growth charts.

$$deviation \sim age +sex+1$$

$$deviation \sim age+1$$

$$deviation \sim sex+1$$

$$deviation \sim1$$

$$deviation \sim0$$

**Equation S 2.** Models for the σ parameter in 22q11DS growth charts.

In the subset of patients with longitudinal imaging (N = 19), we included all available scans, with observations weighted by the inverse of the number of scans per participant to account for repeated measures [28]. An iterative model selection procedure was used to exclude outliers (absolute residuals > 3.5) before reconstructing the final models. The final models yielded two outputs: (a) 22q11DS-specific deviation scores, quantifying how an individual deviates from their cohort norm, and (b) centile lines characterizing this distribution. By combining the SLIP and 22q11DS-adjusted models, we back-transformed the centile lines to generate population growth charts for 22q11DS. Worm plots for global features are shown in **Figure S12**.

#### Sensitivity analyses

##### Controlling for intracranial volume

First, to ensure that observed effects were not driven solely by global volume changes, we repeated the primary analyses while including total intracranial volume (ICV) deviation score as a covariate in the models (**Equation S3**).

$$deviation\sim dx+age_{days}+sex+euler+deviation_{ICV}$$

**Equation S3.** Case-control regression model controlling for intracranial volume.

The main findings remained, although modest changes in the statistical significance and effect direction of some regions were noted (**Table S13** and **Figure S13**). We observed that 37 models were no longer significant but remained directionally consistent with the primary findings, 7 models were no longer significant and the effect had an opposite direction, and 25 models gained significance that were not significant in the primary findings, of which 10 were directionally consistent with the primary findings and 15 were not. No regions were significant in both models with opposing effect directions (i.e., changing from decreased volume in 22q11DS to increased volume relative to intracranial volume). 112 features remained significant and directionally consistent in both models. Similar observations were observed in the ENIGMA cohort (**Table S14** and **Figure S13**).

##### Inclusion of only MPRAGE scans

Second, given the potential for segmentations generated from heterogeneous, clinical data to be less reliable than more traditional research scans, we repeated segmentation and normative growth chart modeling using scans collected with only high resolution (≤ 1.5 mm in all dimensions), T1-weighted scans using the MPRAGE sequence (N = 46, N = 40 for CT).

The main findings remained consistent, although modest reductions in the statistical significance of the results were noted (**Table S15** and **Figure S14**). We observed that 46 models were no longer significant but remained directionally consistent with the primary findings, two models gained significance that were not significant in the primary findings, both of which were directionally consistent with the primary model. The directionality of the effect size did not change in any significant model. 110 features remained significant and directionally consistent in both models. The reduction in the number of significant models is likely partially explained by the reduction in sample size from N = 92 to N = 46.

##### Centiles and non-benchmarked brain features

Third, to assess the reliability of deviation scores as a measure of brain structure, we repeated the primary analysis using (a) brain centiles (i.e., direct outputs from the growth charts prior to transformation into deviation scores) and (b) non-benchmarked brain features (i.e., raw measures of volume, surface area, and thickness without referencing against brain charts).

We observed only minor differences in the models constructed using centile scores compared to deviation scores (**Table S16** and **Figure S15**), with 141 features in the CHOP clinical cohort remaining significant and directionally consistent between the two models. 15 models were no longer significant but remained directionally consistent with the primary findings, one model gained significance that was not significant in the primary findings and was directionally consistent with the primary model. The directionality of the effect size did not change in any significant model. Similar observations were observed in the ENIGMA cohort (**Table S17** and **Figure S15**).

For the non-benchmarked features, we included additional covariates consistent with previous studies of 22q11DS (**Equation S4**). For volumes, we included age, age^2^, sex, and Euler number as covariates [15]. For cortical surface area, we included age, sex, and Euler number as covariates [16]. For cortical thickness, we included age^2^, sex, and Euler number as covariates [16].

$$Volume \sim dx+age_{days}+age_{days}^{2}+sex+euler$$

$$Surface Area \sim dx+age_{days}+sex+euler$$

$$Thickness \sim dx+age_{days}^{2}+sex+euler$$

**Equation S4.** Case-control regression model of non-benchmarked brain features.

We observed only minor differences in the models constructed using non-benchmarked features compared to deviation scores (**Table S18** and **Figure S16**), with 142 features in the CHOP clinical cohort remaining significant and directionally consistent between the two models. 14 models were no longer significant but remained directionally consistent with the primary findings, six models gained significance that was not significant in the primary findings and were directionally consistent with the primary model. The directionality of the effect size did not change in any significant model. Similar observations were observed in the ENIGMA cohort (**Table S19** and **Figure S16**).

##### Inclusion of scans with pathology

Fourth, we reintroduced scans that were determined to have pathology in neuroradiological review that could impact segmentation quality, as long as the segmentation and surface quality of the scan was determined to be sufficient (as assessed via Euler number and SynthSeg+ QC scores). In total, an additional 18 patients were added to the volumetric and surface area analyses (N = 110) and six patients were added to the cortical thickness analysis (N = 46). We observed that three models were no longer significant but remained directionally consistent with the primary findings, and ten models gained significance that were not significant in the primary findings and were directionally consistent with the primary model (**Table S20** and **Figure S17**). The directionality of the effect size did not change in any significant model. 153 features remained significant and directionally consistent in both models.

##### Scanner covariate model

Fifth, while random scanner effects were incorporated into the GAMLSS model and thus deviation scores were calculated while incorporating site-specific effects, we did not further control for the scanner in the main analysis. In this sensitivity analysis, we also employed a covariate for the scanner to model effect sizes in 22q11DS features while accounting for the scanner (**Equation S5**).

$$deviation\sim dx+age_{days}+sex+euler+scanner$$

**Equation S5.** Case-control regression model with fixed effects for MRI scanner.

We observed only minor differences in the models constructed with a covariate for scanner (**Table S21** and **Figure S18**). We observed that 25 models were no longer significant but remained directionally consistent with the primary findings, five models gained significance that were not significant in the primary findings and were directionally consistent with the primary models. The directionality of the effect size did not change in any significant model. 130 features remained significant and directionally consistent in both models. Similar observations were observed in the ENIGMA cohort (**Table S22** and **Figure S18**).

##### Exclusion of young children

Sixth, the inclusion of children below the ages represented in the ENIGMA-22q data could introduce bias. Thus, we limited the CHOP cohort to scans in patients six years or older (N=55, N=31 for CT). We observed only minor differences in the models constructed with a covariate for scanner (**Table S23** and **Figure S19**). We observed that 32 models were no longer significant but remained directionally consistent with the primary findings, five models gained significance that were not significant in the primary findings and were directionally consistent with the primary models. The directionality of the effect size did not change in any significant model. 124 features remained significant and directionally consistent in both models.

### Supplementary Figures

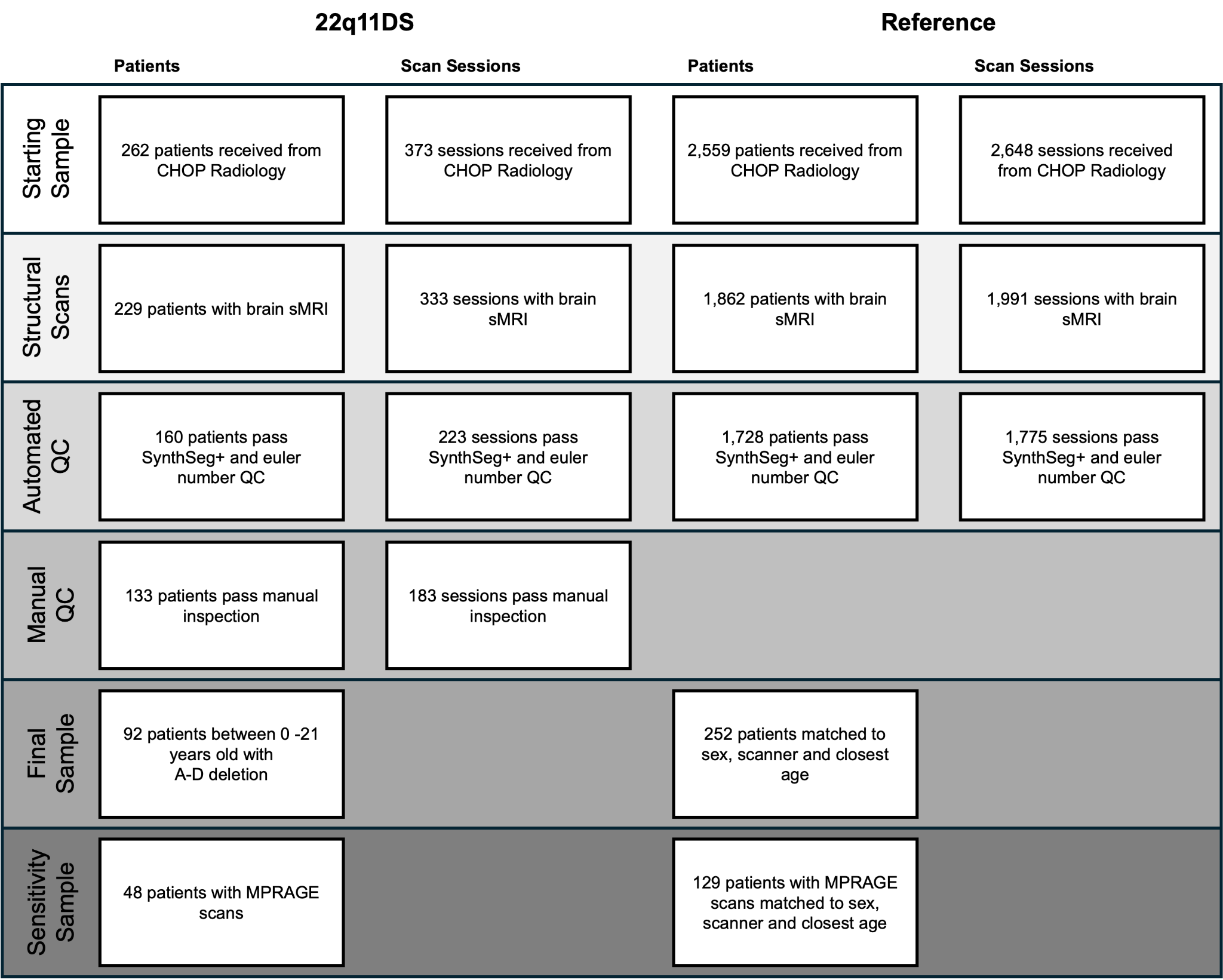

**Figure S1. Sample Selection Flowchart for the Primary Clinical Cohort.** Flowchart of criteria used to derive the final study samples from the initial pool of clinical scans retrieved from the CHOP Radiology department. The left two columns detail the selection process for the 22q11DS cohort, with data presented by the number of unique patients and the number of scan sessions. The right two columns show the corresponding process for the reference control group. The process involved initial sample retrieval, filtering for structural scans (sMRI), and passing both automated (SynthSeg+ and Euler number) and manual quality control steps (for the 22q11DS cohort). The final sample for the primary analysis consisted of 92 patients with the A-D deletion and 252 controls matched for sex, scanner, and age. A subset of these participants with MPRAGE scans comprised the sensitivity analysis cohort, consisting of 48 patients and 129 matched controls.

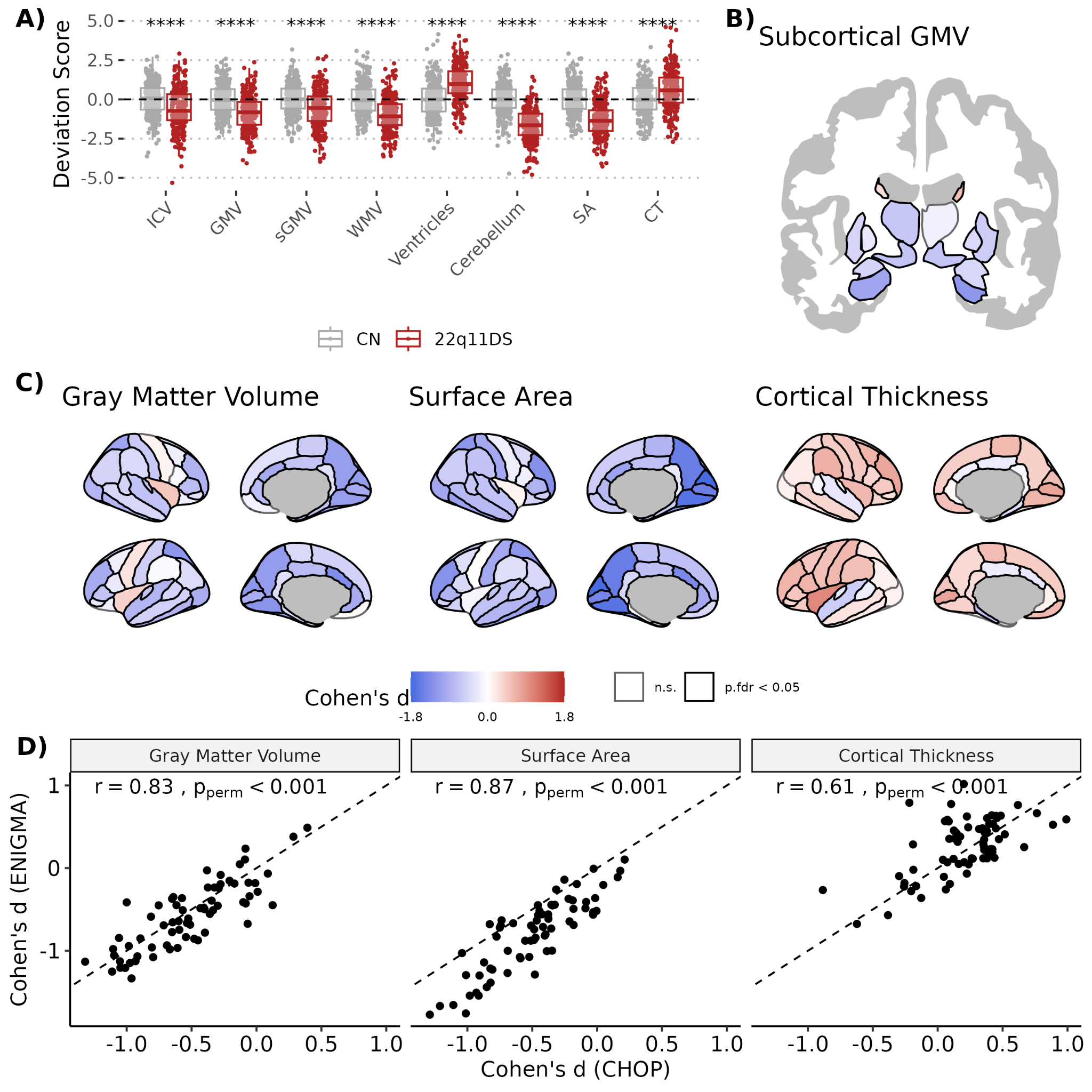

**Figure S2. Effect sizes of 22q11.2 deletion syndrome on brain deviation scores in the ENIGMA-22q cohort. (A)** Boxplots show individual standardized deviation scores for global brain measures in patients with 22q11DS (n=242) and controls in the ENIGMA cohort (n=277). **(B and C)** Brain maps display Cohen’s d effect sizes for regional deviations in **(B)** subcortical gray matter volume (GMV) and **(C)** cortical GMV, surface area (SA), and thickness brain features. Regions with negative deviation scores in 22q11DS (i.e., smaller values in 22q11DS) are shown in blue, and regions with positive scores (i.e., larger values in 22q11DS) are shown in red. Solid black outlines indicate regions with significant case-control differences. **(D)** Scatter plots show the correlation between Cohen’s d effect sizes from the primary CHOP cohort (x-axis) and the independent ENIGMA-22q cohort (y-axis). All deviation scores were derived from normative growth charts predicting brain features based on age, sex, and scanner. P-values were corrected for multiple comparisons using the Benjamini-Hochberg false discovery rate (FDR) procedure. Asterisks indicate the level of statistical significance after correction. **** p < 0.0001, *** p < 0.001, ** p < 0.01 , * < 0.05. Abbreviations: CT, cortical thickness; GMV, gray matter volume; ICV, intracranial volume; n.s., not significant; SA, total SA; sGMV, subcortical GMV; WMV, white matter volume.

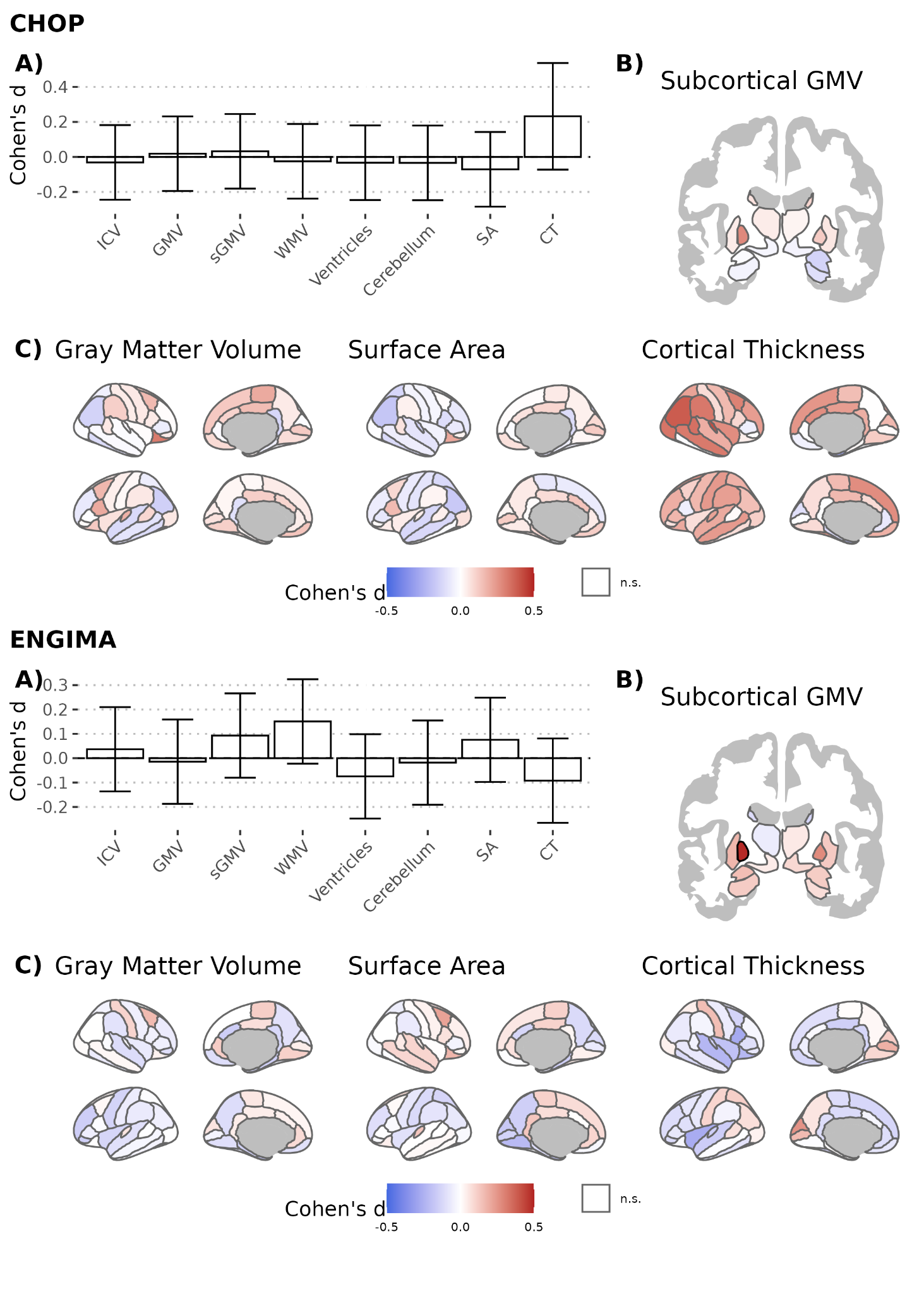

**Figure S3: Age-by-22q11DS interactions of brain deviation scores (A)** Bar charts show Cohen’s d interaction effects for global brain measures with age in patients with 22q11DS (N CHOP = 92, N ENIGMA-22q = 242) and controls (N CHOP = 252, N ENIGMA-22q = 277). Error bars represent 95% confidence intervals **(B and C)** Brain maps display Cohen’s d effect sizes for regional deviations and age in **(B)** subcortical gray matter volume (GMV) and **(C)** cortical GMV, surface area (SA), and thickness brain features. Negative effect sizes indicate decreasing deviation scores with increasing age. Solid black outlines indicate regions with significant case-control differences. All deviation scores were derived from normative growth charts predicting brain features based on age, sex, and scanner. P-values were corrected for multiple comparisons using the Benjamini-Hochberg false discovery rate (FDR) procedure. Asterisks indicate the level of statistical significance after correction. **** p < 0.0001, *** p < 0.001, ** p < 0.01 , * < 0.05. Abbreviations: CT, cortical thickness; GMV, gray matter volume; ICV, intracranial volume; n.s., not significant; SA, total SA; sGMV, subcortical GMV; WMV, white matter volume.

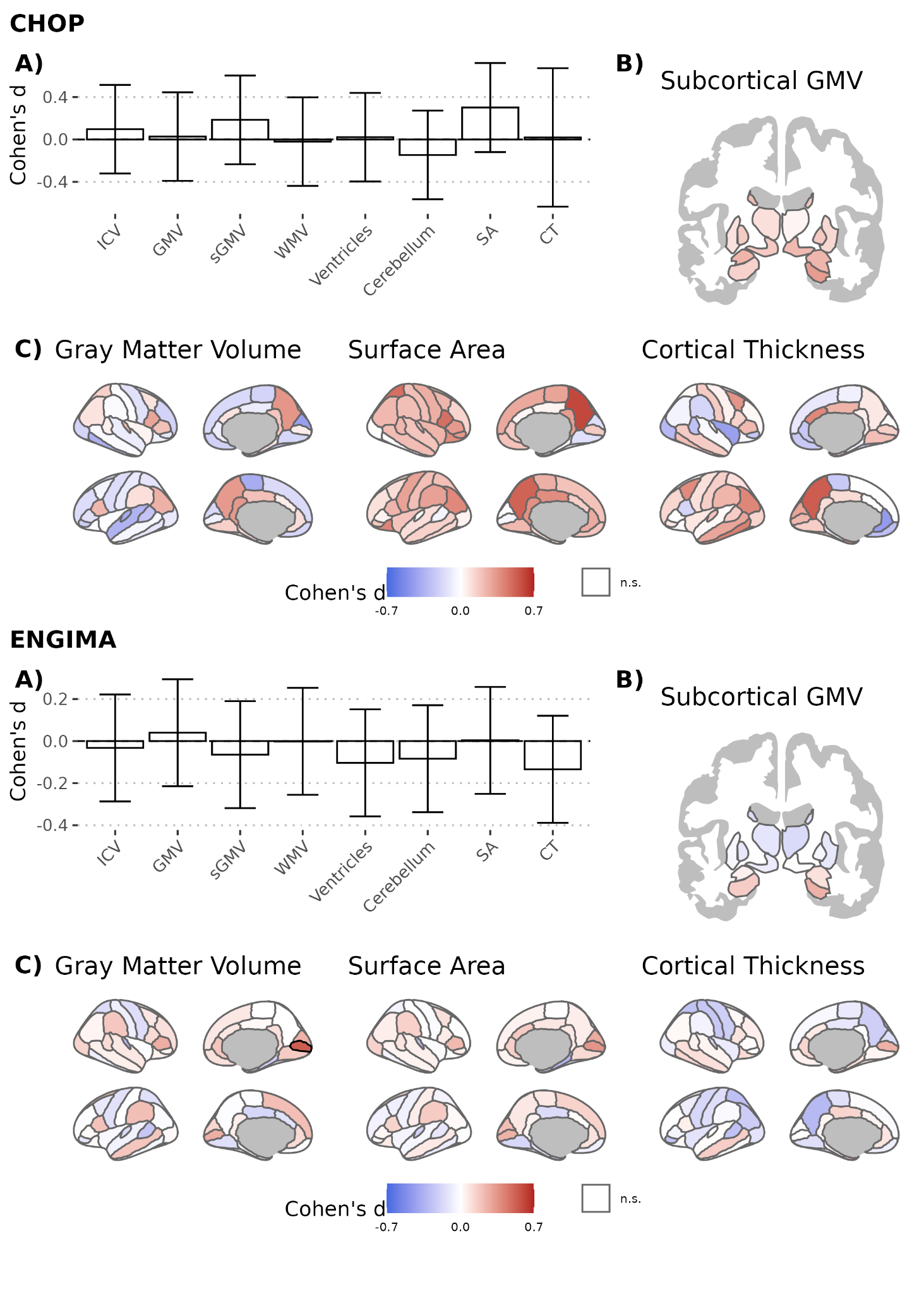

**Figure S4: Sex-by-dx interactions of brain deviation scores. (A)** Bar charts show Cohen’s d interaction effects for global brain measures with sex in patients with 22q11DS (N CHOP = 92, N ENIGMA-22q = 242) and controls (N CHOP = 252, N ENIGMA-22q = 277). Error bars represent 95% confidence intervals **(B and C)** Brain maps display Cohen’s d effect sizes for regional deviations and sex in **(B)** subcortical gray matter volume (GMV) and **(C)** cortical GMV, surface area (SA), and thickness brain features. Negative effect sizes indicate higher deviation sizes in females compared to males. Solid black outlines indicate regions with significant case-control differences. All deviation scores were derived from normative growth charts predicting brain features based on age, sex, and scanner. P-values were corrected for multiple comparisons using the Benjamini-Hochberg false discovery rate (FDR) procedure. Asterisks indicate the level of statistical significance after correction. **** p < 0.0001, *** p < 0.001, ** p < 0.01 , * < 0.05. Abbreviations: CT, cortical thickness; GMV, gray matter volume; ICV, intracranial volume; n.s., not significant; SA, total SA; sGMV, subcortical GMV; WMV, white matter volume.

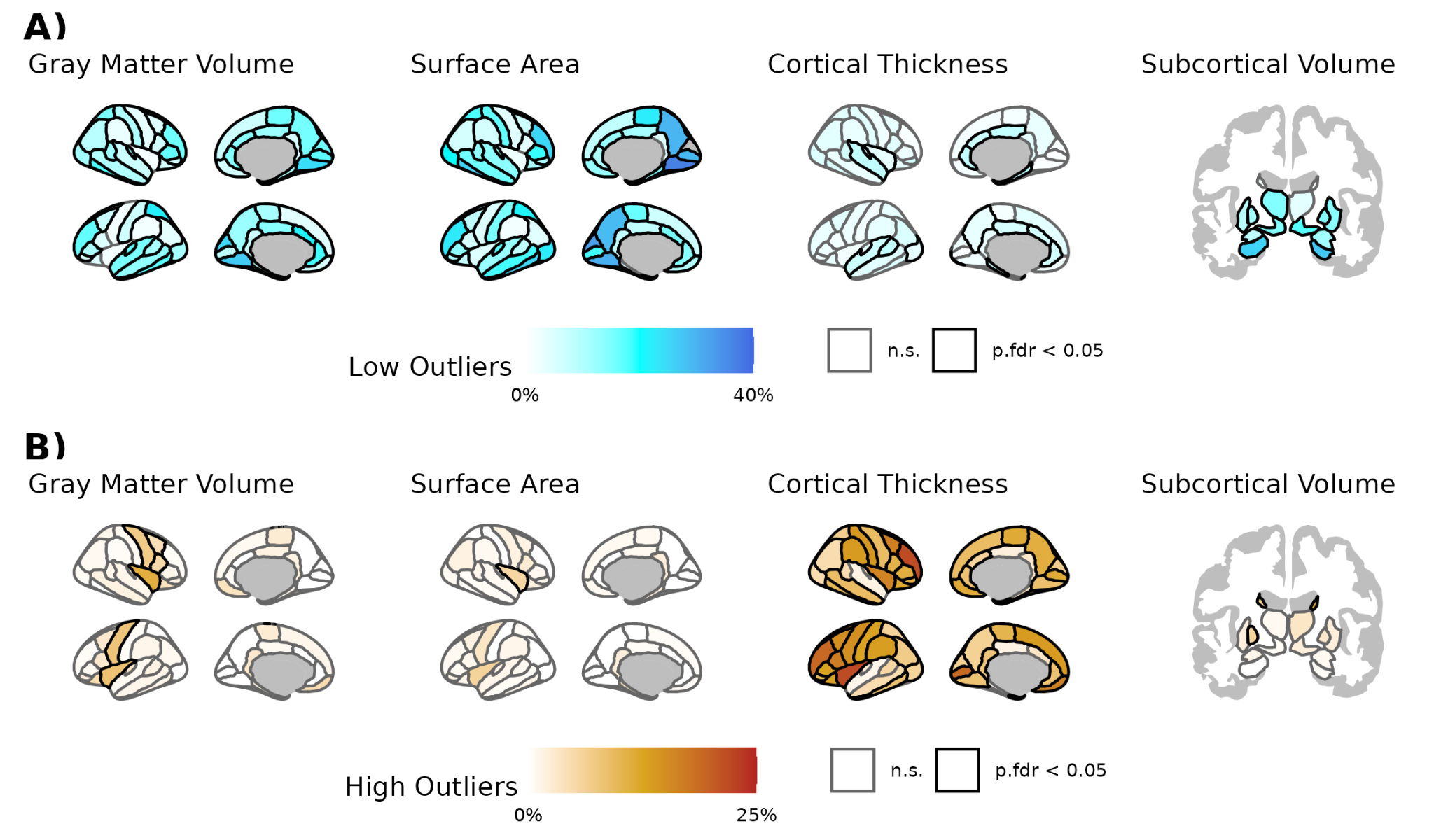

**Figure S5. Extreme atypicality of imaging phenotypes in the ENIGMA-22q cohort.** Brain maps showing the percentage of patients with atypically low scores **(A)** and atypically high scores **(B)** for each brain feature in the ENIGMA-22q cohort. Atypically low scores were defined as falling below the 2.5th percentile of the reference distribution, while atypically high scores were defined as above the 97.5th percentile. The statistical significance of regional enrichment for extreme phenotypes (indicated by solid black outlines) was assessed using a permutation test of case-control diagnoses (1,000 permutations). P-values were corrected for multiple comparisons using the Benjamini-Hochberg false discovery rate (FDR) procedure. n.s. indicates not significant.

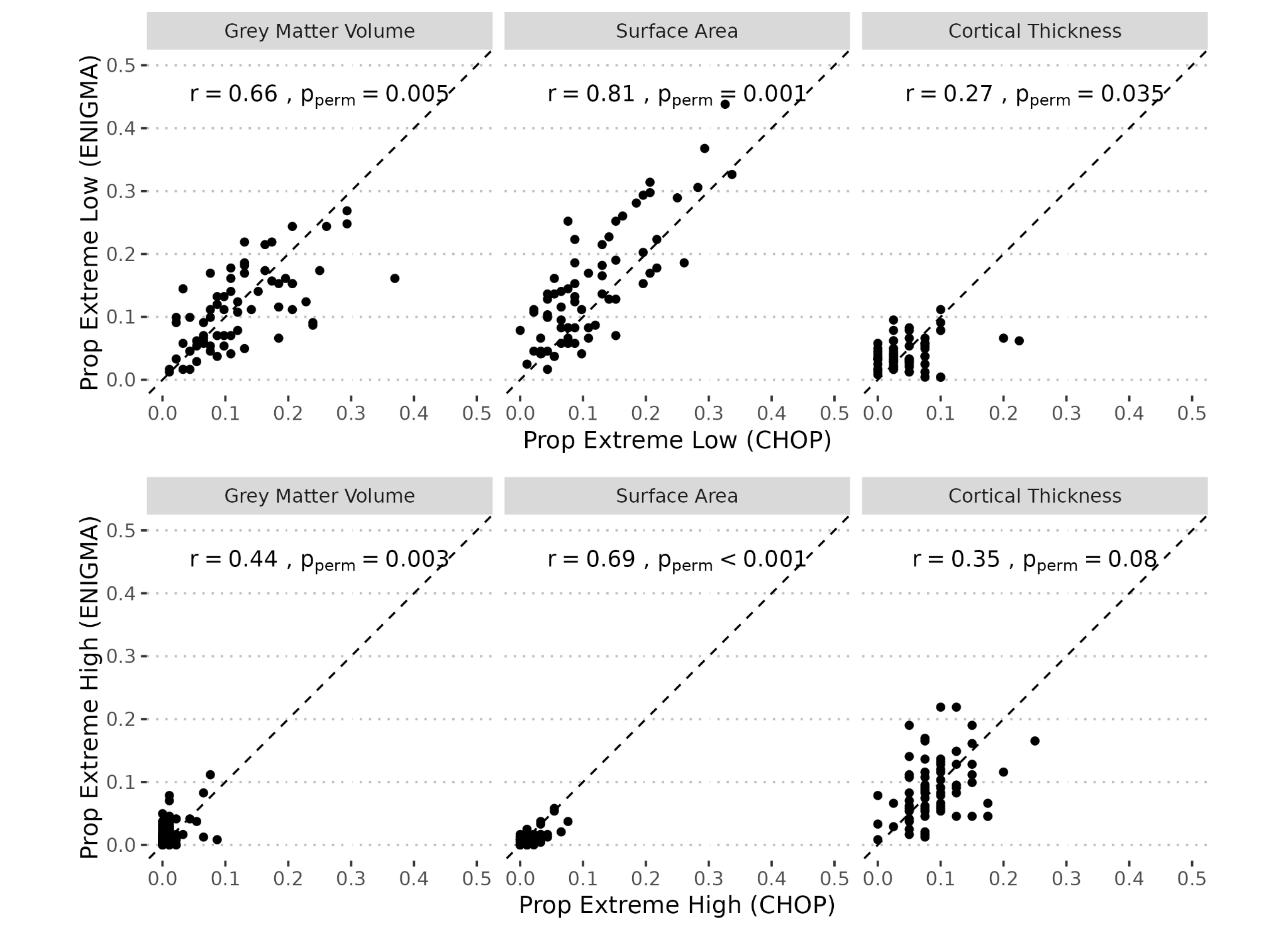

**Figure S6. Correlations between extreme outlier proportions in the CHOP and ENIGMA-22q cohorts.** Scatter plots comparing the proportion of extreme brain atypicality in between the primary clinical (CHOP) cohort and the independent research (ENIGMA-22q) cohort. Each point represents a single cortical region. The x-axis shows the proportion of individuals in the CHOP cohort with an extreme value for that region, while the y-axis shows the corresponding proportion in the ENIGMA-22q cohort. Extreme low proportions (individuals with measures at or below the 2.5th percentile) are shown on the top row, while the bottom row shows the extreme high proportions (individuals with measures at or above the 97.5th percentile). Plots are shown for gray matter volume, surface area, and cortical thickness, with the Pearson correlation coefficient (r) and a permutation-based p-value provided for each set of measures.

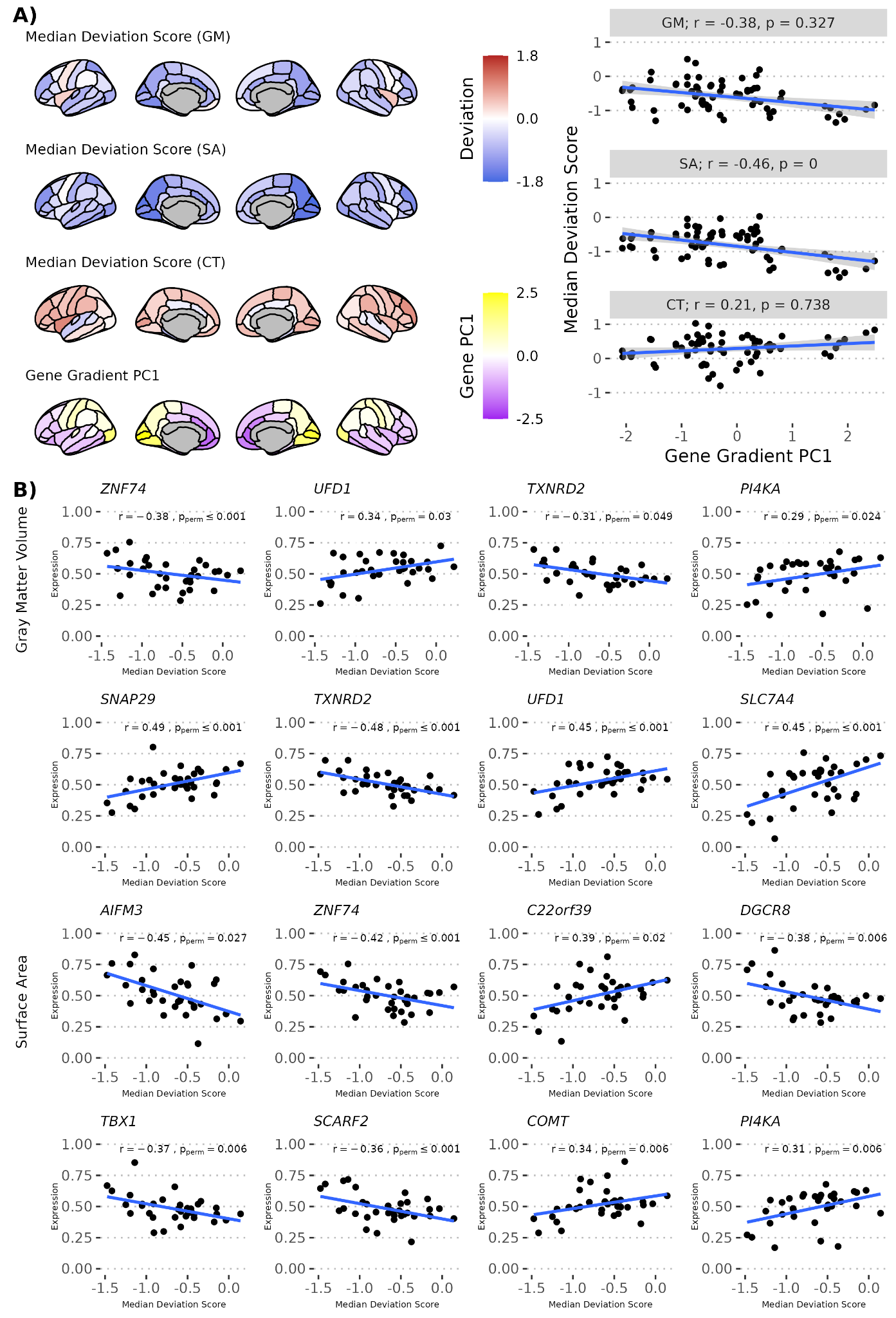

**Figure S7. Spatial correlation between brain deviations and gene expression in the ENIGMA-22q cohort (A)** The spatial maps of median deviation scores in the ENIGMA-22q cohort for gray matter volume (GMV), surface area (SA), and cortical thickness (CT) compared to the first principal component (PC1) of brain-wide gene expression from the Allen Human Brain Atlas. Scatter plots show a negative correlation between PC1 and deviation maps for SA (r=−0.46, p_perm_<0.001). **(B)** Shown are the twelve 22q11DS genes that were significantly correlated with deviation maps after multiple comparisons correction. Significance was assessed by comparing the difference in correlation coefficients of cases and controls to a null distribution across 1,000 permutations. Abbreviations: CT, cortical thickness; GMV, gray matter volume; PC1, first principal component; SA, surface area.

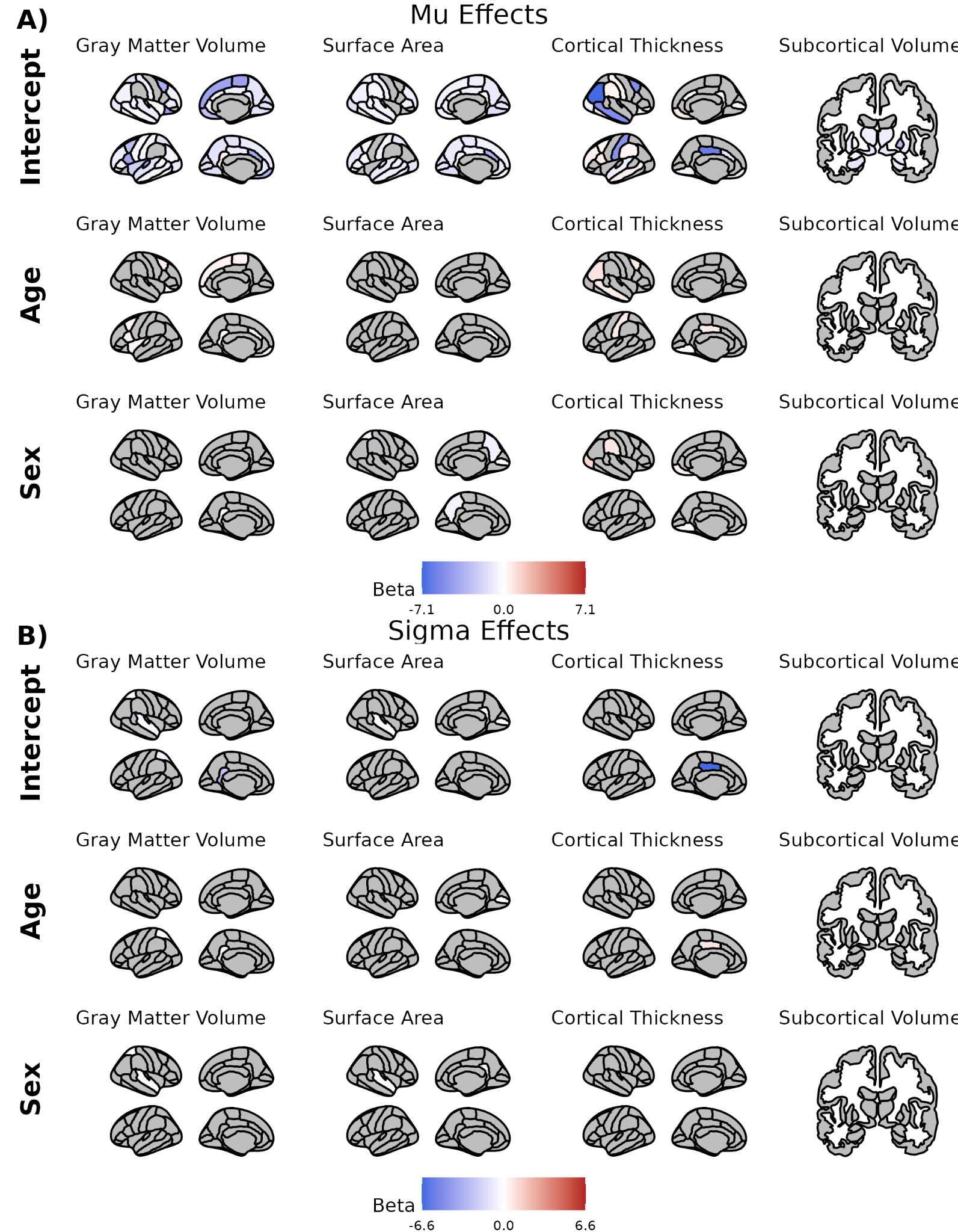

**Figure S8. Regional LMSz coefficients for 22q11DS-adjusted growth charts.** Brain maps of the beta coefficients from the regional LMSz models, illustrating the effects of age, sex and the model intercept on the 22q11DS-specific growth chart parameters. The top rows show the model coefficients for the model’s mean parameter (µ), which represents the mean deviation score within the 22q11DS cohort. The bottom section shows the model coefficients for the model’s scale parameter (σ), representing the variance of the deviation scores. Within each section, the rows display the effects of age, sex and intercept, while the columns correspond to different brain features. The color of each brain region indicates the value of the beta coefficient, with red indicating a positive association (e.g., the deviation score increases with age) and blue indicating a negative association. Grey regions indicate that the parameter was not included in the final model.

**Figure S9: Examples of excluded pathology in 22q11DS.** Representative T1-weighted MR images from three different individuals with 22q11DS who were excluded from the primary analysis are shown. These scans were excluded due to the presence of significant structural pathology that could confound the automated segmentation of brain volumes. **(A)** Sagittal view showing the presence of a Chiari malformation. **(B)** Axial view showing evidence of extensive polymicrogyria (atypically small and numerous brain folds). **(C)** Axial view showing severe ventriculomegaly (enlarged ventricles).

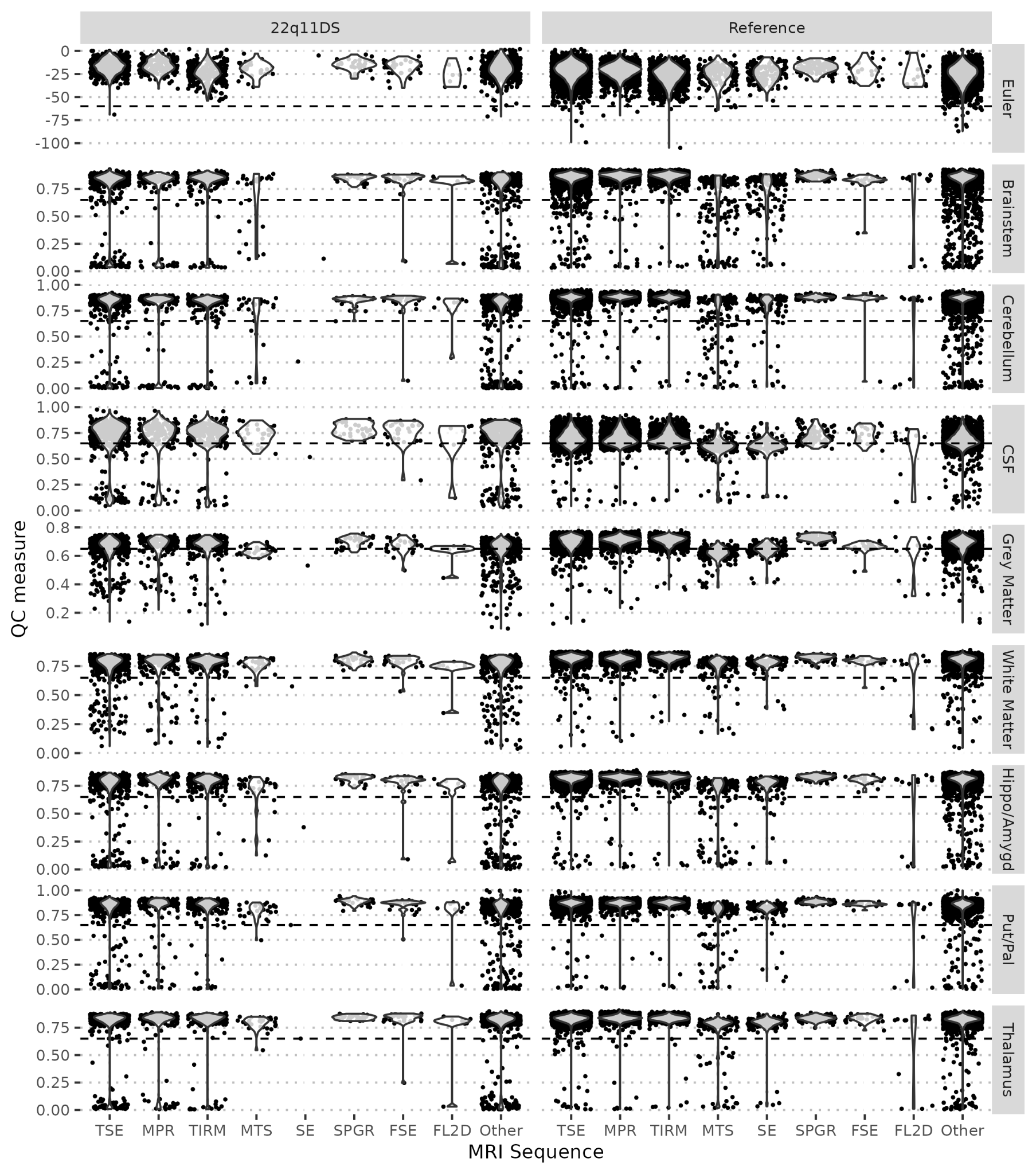

**Figure S10: Automated QC measures by scan sequence.** Violin plots showing the distribution of the mean Euler number and SynthSeg+ Dice scores, automated metrics of image quality and segmentation, for each MRI scan separated by scan sequence. Data are shown for both the 22q11DS cohort (left) and the reference cohort (right). The dashed horizontal line indicates the QC exclusion threshold (-60 for Euler, 0.65 for SynthSeg+ scores). Exclusion rates varied by scan sequence (least excluded SPGR: 16%, most excluded MTS: 69%). Abbreviations: TSE, Turbo spin echo; MPRAGE, Magnetization-Prepared Rapid Acquisition Gradient Echo; TIRM, Turbo inversion recovery magnitude; SPGR, spoil gradient recalled echo; MTS, Magnetization Transfer Saturation; GE, Gradient Echo

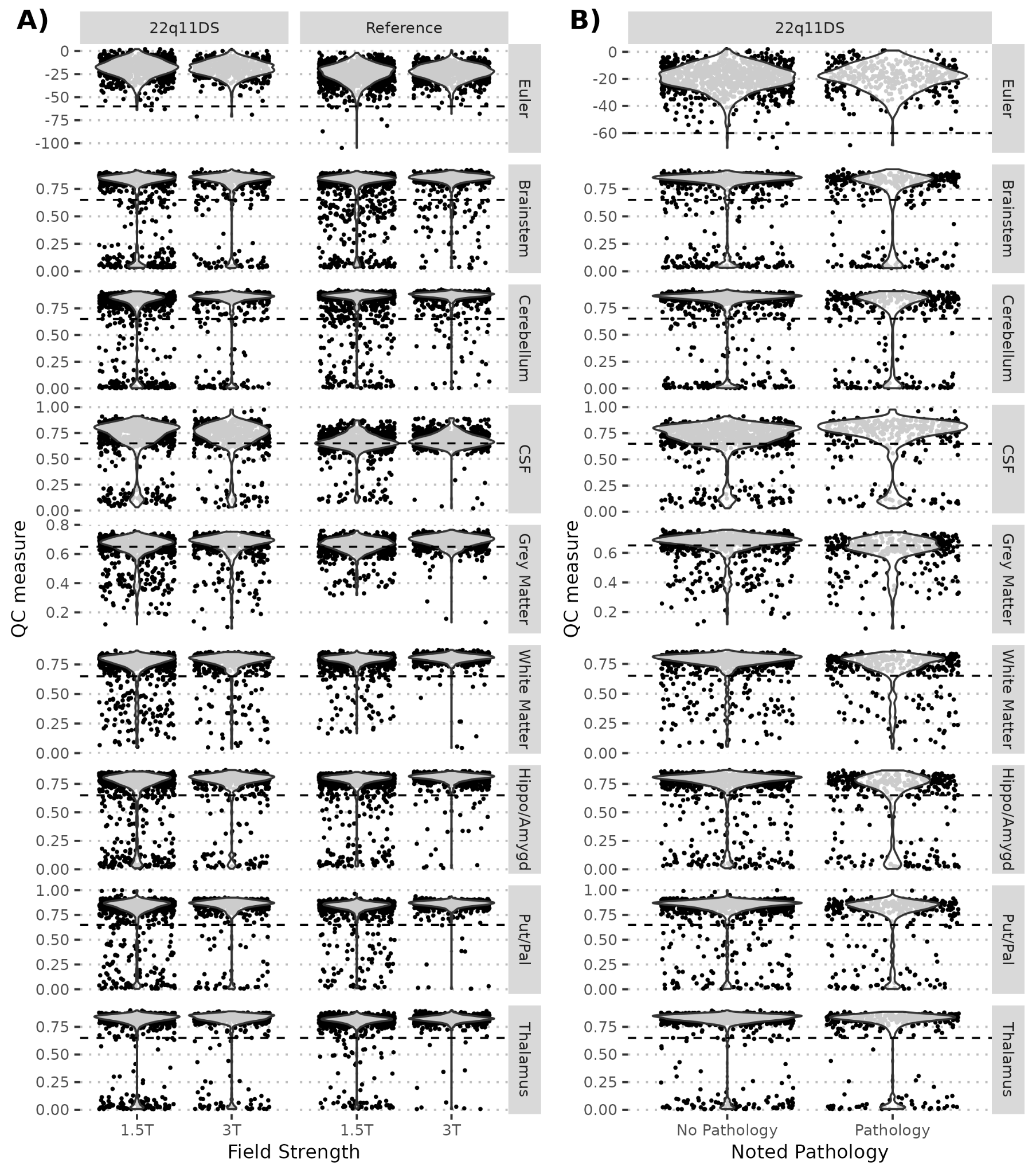

**Figure S11: Automated QC measures by field strength and pathology.** Violin plots showing the distribution of the mean Euler number and SynthSeg+ Dice scores, automated metrics of image quality and segmentation, for each MRI scan separated by **(A)** field strength and **(B)** pathology noted by a neuroradiologist. Data are shown for both the 22q11DS cohort (left) and the reference cohort (right) for field strength. The dashed horizontal line indicates the QC exclusion threshold (-60 for Euler, 0.65 for SynthSeg+ scores). Exclusion rates varied by field strength (1.5T: 44% excluded, 3.0T: 33% excluded) and pathology (pathology: 58% excluded, no pathology: 33% excluded).

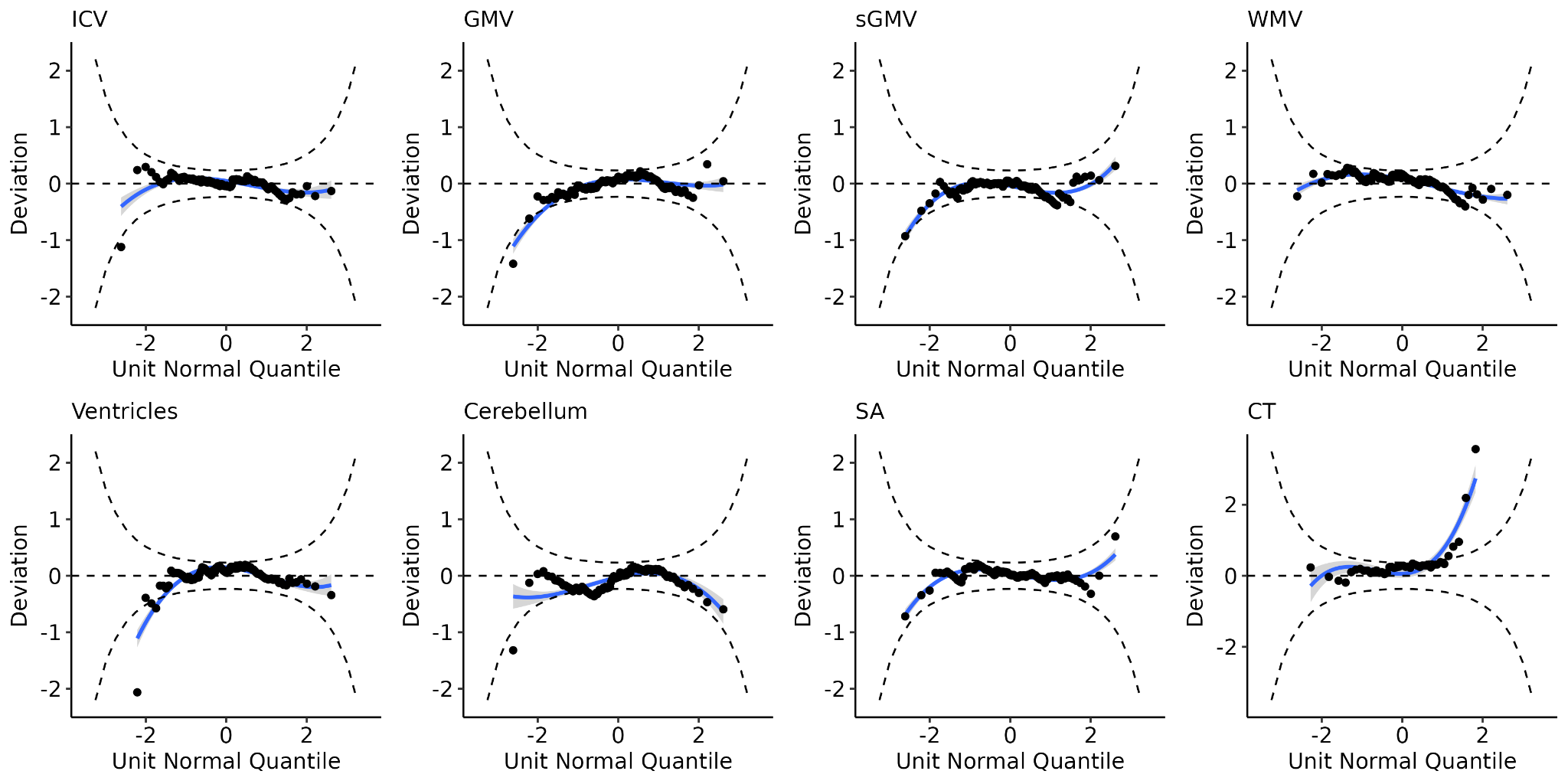

**Figure S12: Worm plots for global 22q11DS-adjusted growth charts.** Worm plots (detrended Q-Q plots) used to visually assess the goodness-of-fit of the 22q11DS-specific GAMLSS models for each global brain measure. Each plot shows the deviations of quantile residuals from the model plotted against their expected quantiles from a standard normal distribution. The solid blue line represents the median (50th centile), while the dashed lines represent the outer centiles of the distribution. For a perfect model fit, the smoothed centile lines should be horizontal and centered at a deviation of zero. The plots indicate a good model fit, showing that the resulting adjusted deviation scores are approximately normally distributed. Abbreviations: CT, cortical thickness; GMV, gray matter volume; ICV, intracranial volume; SA, total surface area; sGMV, subcortical gray matter volume; WMV, white matter volume.

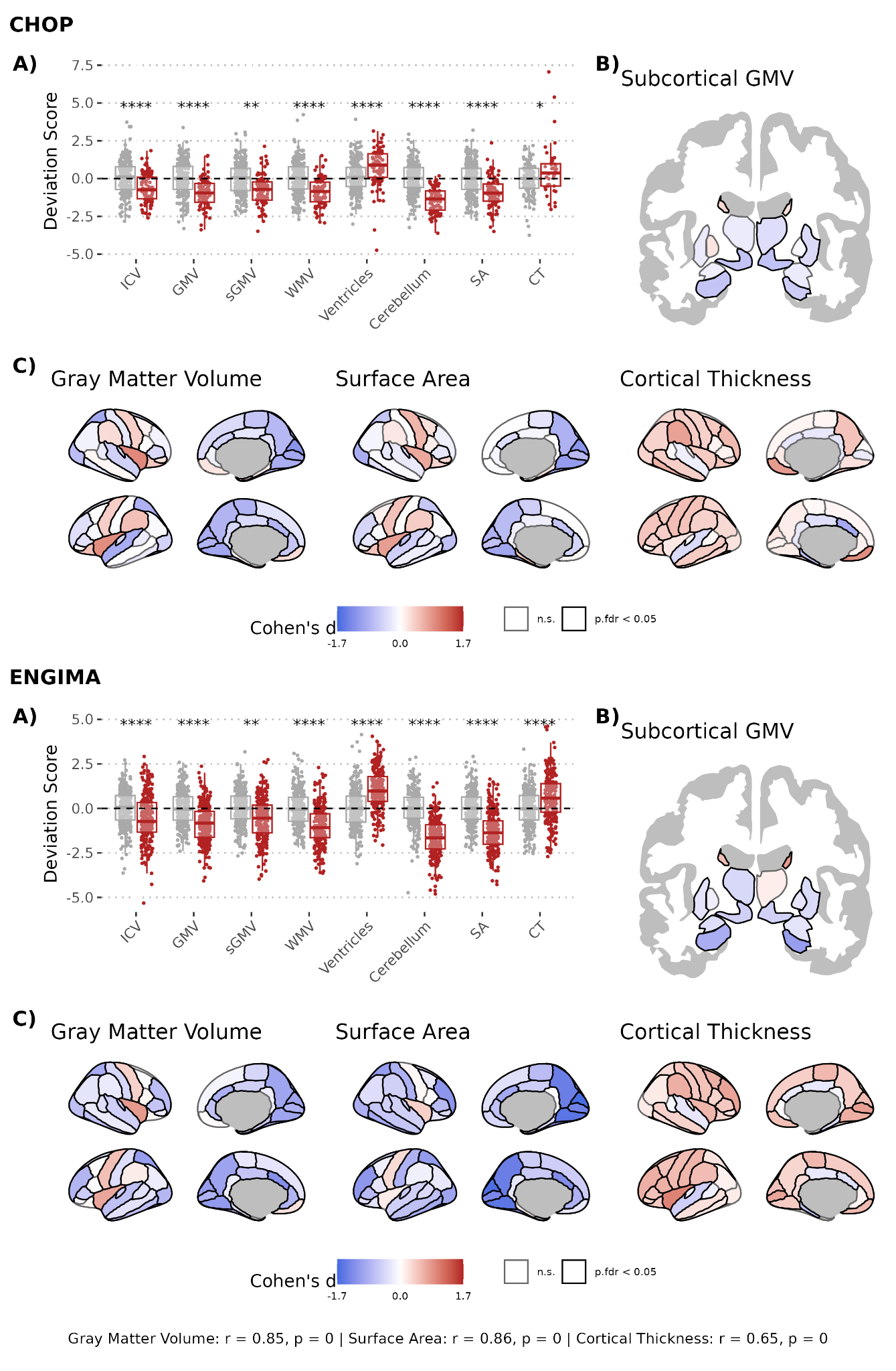

**Figure S13. Sensitivity analysis: Controlling for intracranial volume.** Cohen’s d effect sizes for the CHOP clinical primary cohort (left panel) and the ENIGMA-22q research cohort (right panel). Compared to the primary analysis, a covariate for the intracranial volume deviation score was included to account for global brain size. **(A)** Box plots show individual standardized deviation scores for global brain measures in patients with 22q11DS (N CHOP = 92, N ENIGMA-22q = 242) and controls (N CHOP = 252, N ENIGMA-22q = 277). **(B and C)** Brain maps display Cohen’s d effect sizes for regional deviations in **(**B**)** subcortical gray matter volume (GMV) and **(C)** cortical GMV, surface area (SA), and thickness brain features. Regions with negative deviation scores in 22q11DS (i.e., smaller values in 22q11DS) are shown in blue, and regions with positive scores (i.e., larger values in 22q11DS) are shown in red. Solid black outlines indicate regions with significant case-control differences. **(D)** Scatter plots show the correlation between Cohen’s d effect sizes from the primary CHOP cohort (x-axis) and the independent ENIGMA-22q cohort (y-axis). All deviation scores were derived from normative growth charts predicting brain features based on age, sex, and scanner. P-values were corrected for multiple comparisons using the Benjamini-Hochberg false discovery rate (FDR) procedure. Asterisks indicate the level of statistical significance after correction. **** p < 0.0001, *** p < 0.001, ** p < 0.01 , * < 0.05. Abbreviations: CT, cortical thickness; GMV, gray matter volume; ICV, intracranial volume; n.s., not significant; SA, total SA; sGMV, subcortical GMV; WMV, white matter volume.

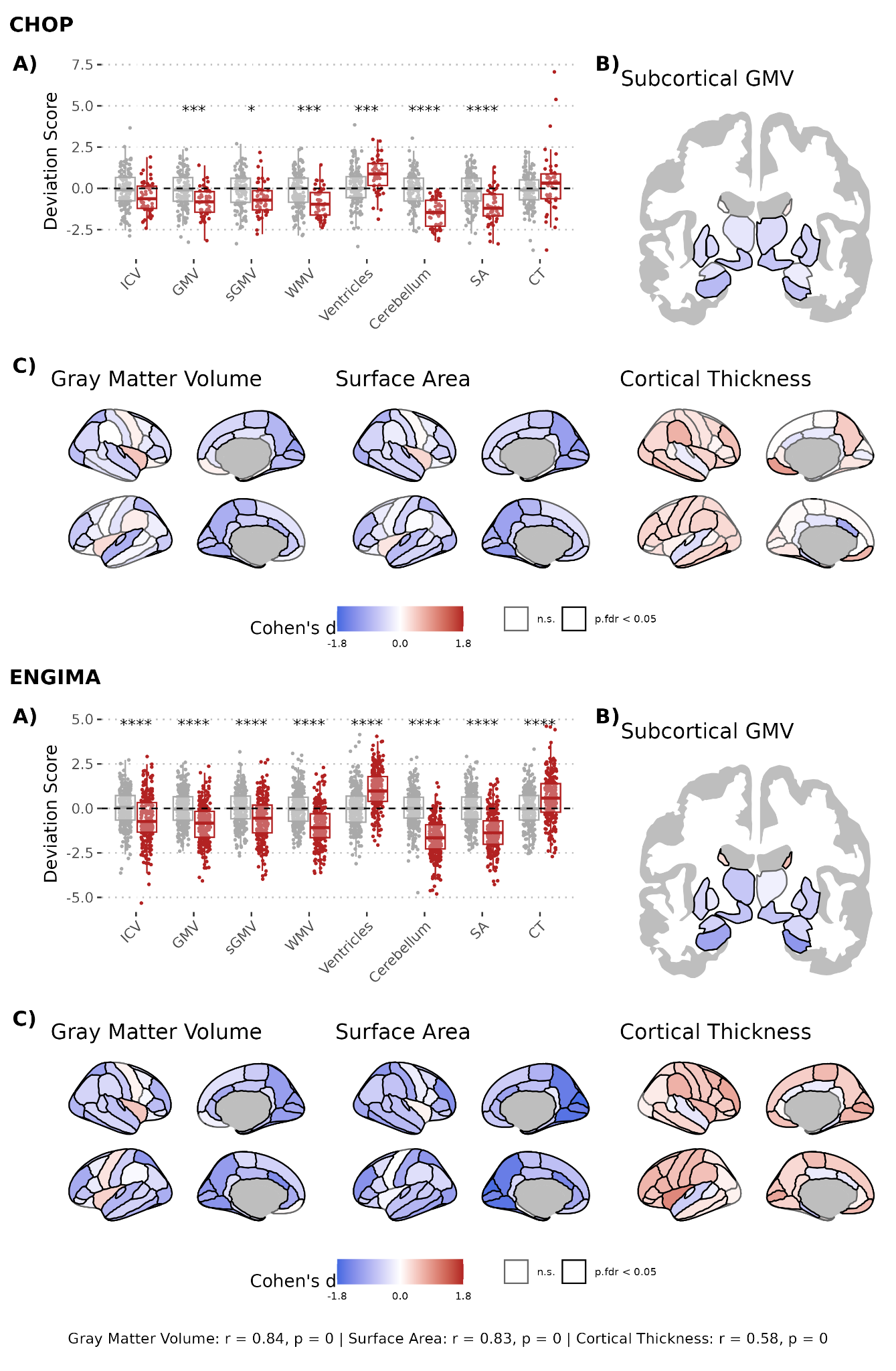

**Figure S14. Sensitivity analysis: MPRAGE clinical scans.** Cohen’s d effect sizes for the CHOP clinical primary cohort (left panel) and the ENIGMA-22q research cohort (right panel). Compared to the primary analysis, scans were restricted to high resolution (≤ 1.5 mm in all dimensions) scans using the magnetization-prepared rapid acquisition gradient-echo (MPRAGE) sequence. This subset of clinical scans was then compared to the complete ENIGMA-22q cohort. **(A)** Box plot show individual standardized deviation scores for global brain measures in patients with 22q11DS (N CHOP = 48, N ENIGMA-22q = 242) and controls (N CHOP = 129, N ENIGMA-22q = 277). **(B and C)** Brain maps display Cohen’s d effect sizes for regional deviations in **(B)** subcortical gray matter volume (GMV) and **(C)** cortical GMV, surface area (SA), and thickness brain features. Regions with negative deviation scores in 22q11DS (i.e., smaller values in 22q11DS) are shown in blue, and regions with positive scores (i.e., larger values in 22q11DS) are shown in red. Solid black outlines indicate regions with significant case-control differences. **(D)** Scatter plots show the correlation between Cohen’s d effect sizes from the primary CHOP cohort (x-axis) and the independent ENIGMA-22q cohort (y-axis). All deviation scores were derived from normative growth charts predicting brain features based on age, sex, and scanner. P-values were corrected for multiple comparisons using the Benjamini-Hochberg false discovery rate (FDR) procedure. Asterisks indicate the level of statistical significance after correction. **** p < 0.0001, *** p < 0.001, ** p < 0.01 , * < 0.05. Abbreviations: CT, cortical thickness; GMV, gray matter volume; ICV, intracranial volume; n.s., not significant; SA, total SA; sGMV, subcortical GMV; WMV, white matter volume.

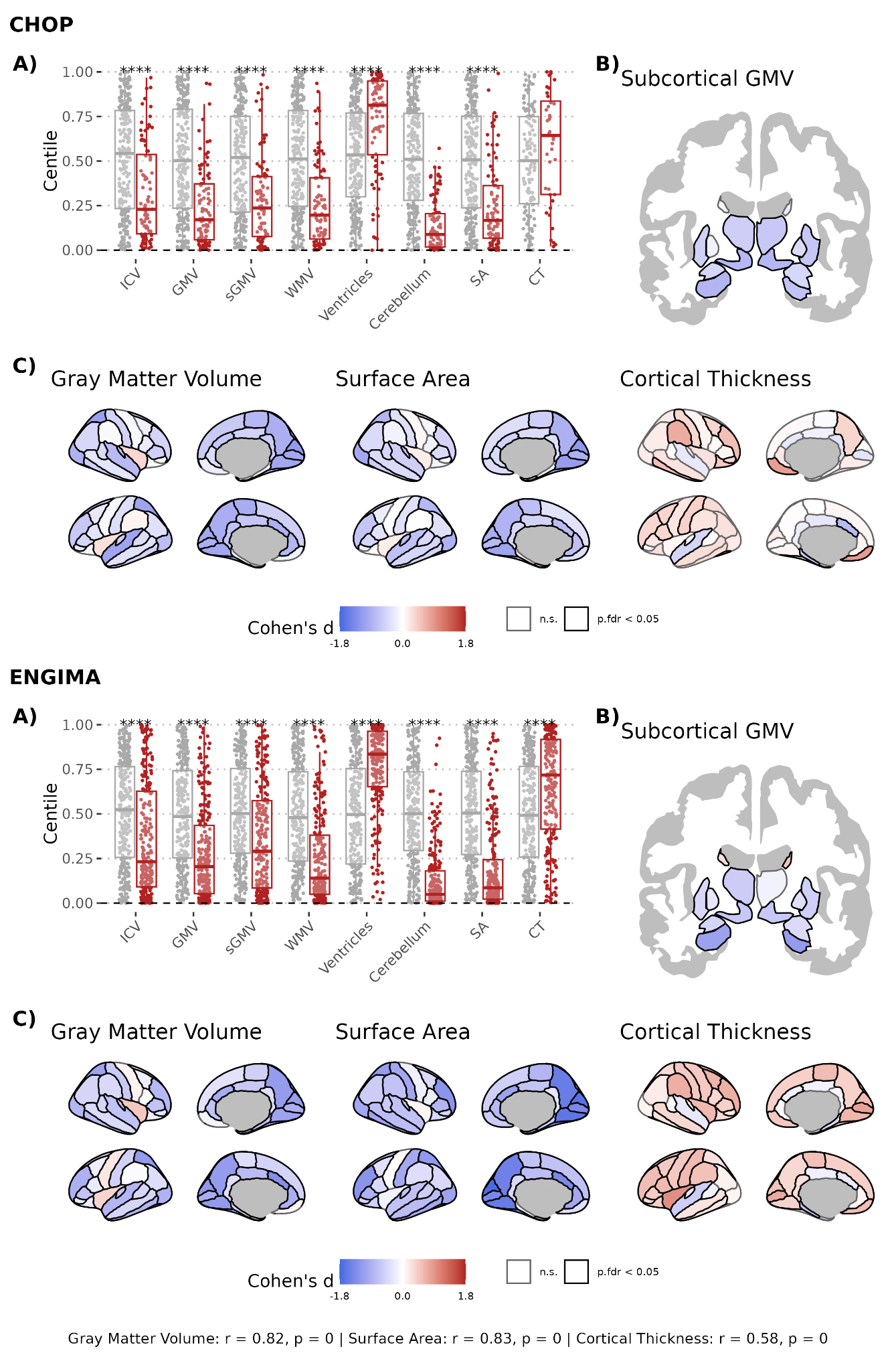

**Figure S15. Sensitivity analysis: Brain centiles.** Cohen’s d effect sizes for the CHOP clinical primary cohort (left panel) and the ENIGMA-22q research cohort (right panel). Compared to the primary analysis, the centile of the brain region was used in place of the brain deviation score. A brain centile of 0 indicates that the measure is among the lowest possible values, a brain centile of 1 indicates that the measure is among the highest possible values, and a brain centile of 0.5 indicates that the measure is at the expected level for a given age and sex. **(A)** Box plot show individual standardized deviation scores for global brain measures in patients with 22q11DS (N CHOP = 92, N ENIGMA-22q = 242) and controls (N CHOP = 252, N ENIGMA-22q = 277). **(B and C)** Brain maps display Cohen’s d effect sizes for regional deviations in **(B)** subcortical gray matter volume (GMV) and **(C)** cortical GMV, surface area (SA), and thickness brain features. Regions with negative deviation scores in 22q11DS (i.e., smaller values in 22q11DS) are shown in blue, and regions with positive scores (i.e., larger values in 22q11DS) are shown in red. Solid black outlines indicate regions with significant case-control differences. (D) Scatter plots show the correlation between Cohen’s d effect sizes from the primary CHOP cohort (x-axis) and the independent ENIGMA-22q cohort (y-axis). All deviation scores were derived from normative growth charts predicting brain features based on age, sex, and scanner. P-values were corrected for multiple comparisons using the Benjamini-Hochberg false discovery rate (FDR) procedure. Asterisks indicate the level of statistical significance after correction. **** p < 0.0001, *** p < 0.001, ** p < 0.01 , * < 0.05. Abbreviations: CT, cortical thickness; GMV, gray matter volume; ICV, intracranial volume; n.s., not significant; SA, total SA; sGMV, subcortical GMV; WMV, white matter volume.

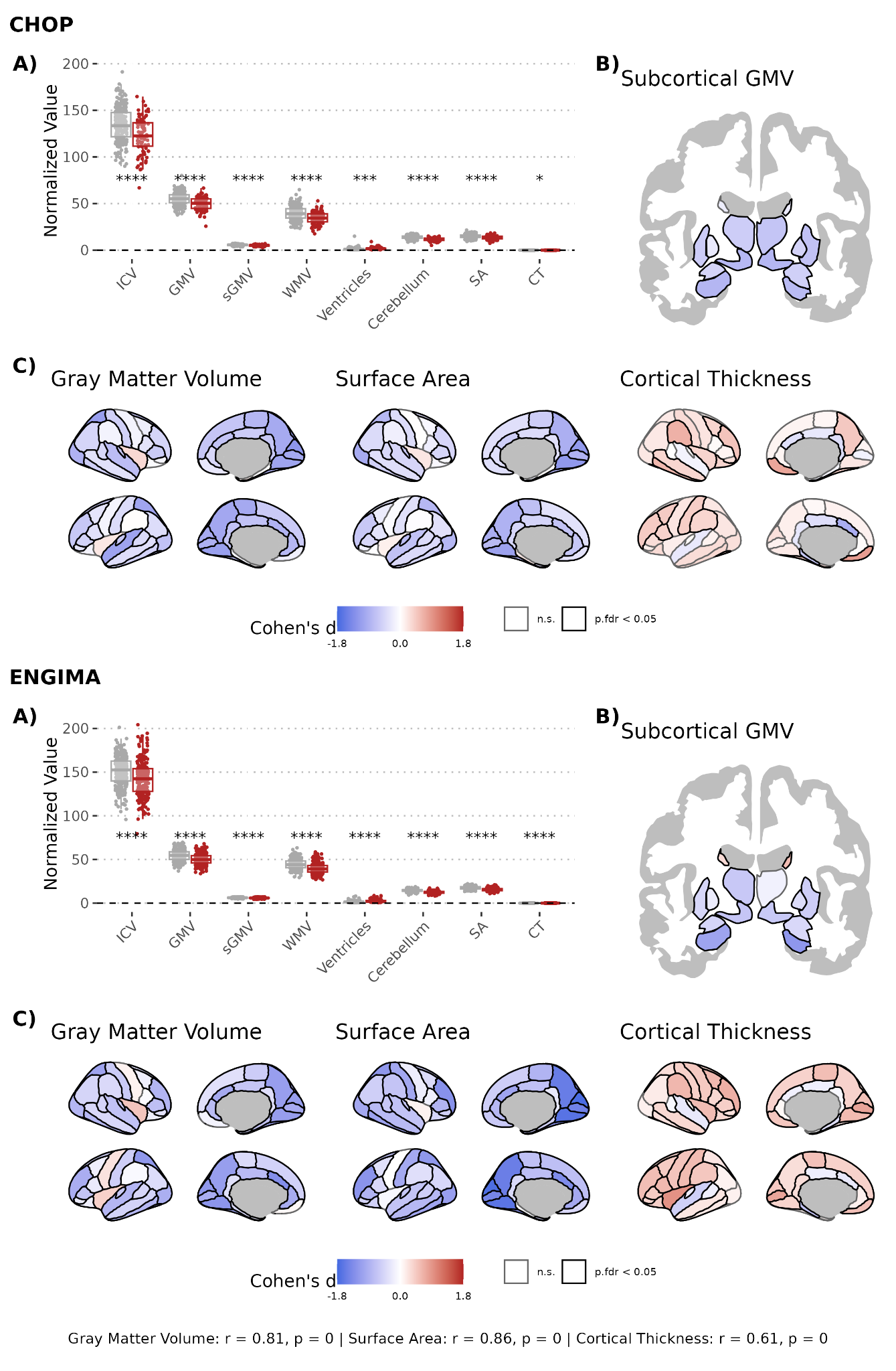

**Figure S16. Sensitivity analysis: Non-normative brain phenotypes.** Cohen’s d effect sizes for the CHOP clinical primary cohort (left panel) and the ENIGMA-22q research cohort (right panel). Compared to the primary analysis, the non-normative measures of volume, surface area, and cortical thickness were used in place of the brain deviation score. **(A)** Box plot show individual standardized deviation scores for global brain measures in patients with 22q11DS (N CHOP = 92, N ENIGMA-22q = 242) and controls (N CHOP = 252, N ENIGMA-22q = 277). **(B and C)** Brain maps display Cohen’s d effect sizes for regional deviations in **(B)** subcortical gray matter volume (GMV) and **(C)** cortical GMV, surface area (SA), and thickness brain features. Regions with negative deviation scores in 22q11DS (i.e., smaller values in 22q11DS) are shown in blue, and regions with positive scores (i.e., larger values in 22q11DS) are shown in red. Solid black outlines indicate regions with significant case-control differences. **(D)** Scatter plots show the correlation between Cohen’s d effect sizes from the primary CHOP cohort (x-axis) and the independent ENIGMA-22q cohort (y-axis). All deviation scores were derived from normative growth charts predicting brain features based on age, sex, and scanner. P-values were corrected for multiple comparisons using the Benjamini-Hochberg false discovery rate (FDR) procedure. Asterisks indicate the level of statistical significance after correction. **** p < 0.0001, *** p < 0.001, ** p < 0.01 , * < 0.05. Abbreviations: CT, cortical thickness; GMV, gray matter volume; ICV, intracranial volume; n.s., not significant; SA, total SA; sGMV, subcortical GMV; WMV, white matter volume.

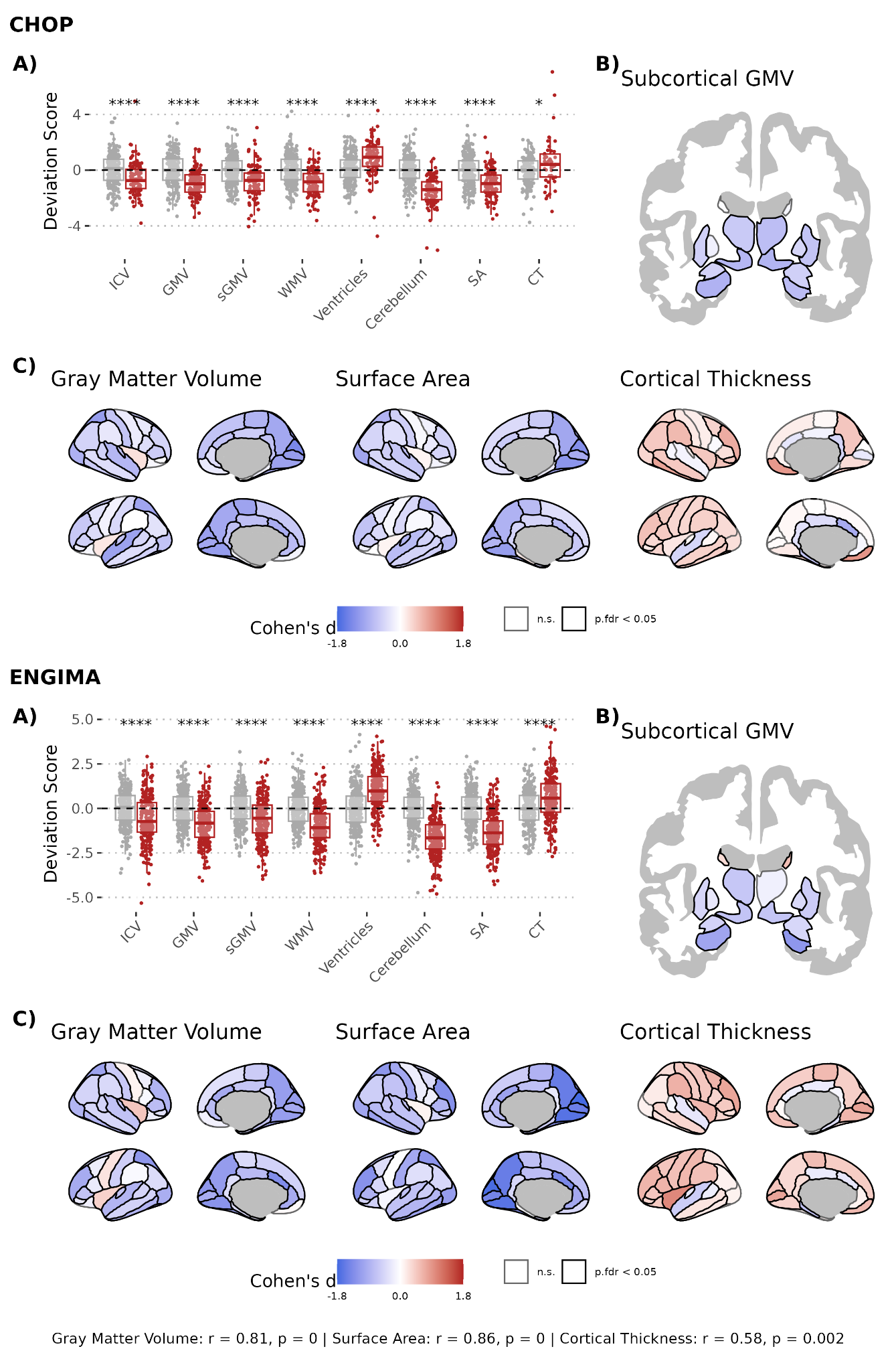

**Figure S17. Sensitivity analysis: Including individuals with identified brain pathologies.** Cohen’s d effect sizes for the CHOP clinical primary cohort (left panel) and the ENIGMA-22q research cohort (right panel). Compared to the primary analysis, additional scans were included that had pathologies identified by a neuroradiologist that may impact segmentation quality (N = 18). This expanded dataset of clinical scans was then compared to complete ENIGMA-22q cohort. **(A)** Box plot show individual standardized deviation scores for global brain measures in patients with 22q11DS (N CHOP = 48, N ENIGMA-22q = 242) and controls (N CHOP = 129, N ENIGMA-22q = 277). **(B and C)** Brain maps display Cohen’s d effect sizes for regional deviations in **(B)** subcortical gray matter volume (GMV) and **(C)** cortical GMV, surface area (SA), and thickness brain features. Regions with negative deviation scores in 22q11DS (i.e., smaller values in 22q11DS) are shown in blue, and regions with positive scores (i.e., larger values in 22q11DS) are shown in red. Solid black outlines indicate regions with significant case-control differences. **(D)** Scatter plots show the correlation between Cohen’s d effect sizes from the primary CHOP cohort (x-axis) and the independent ENIGMA-22q cohort (y-axis). All deviation scores were derived from normative growth charts predicting brain features based on age, sex, and scanner. P-values were corrected for multiple comparisons using the Benjamini-Hochberg false discovery rate (FDR) procedure. Asterisks indicate the level of statistical significance after correction. **** p < 0.0001, *** p < 0.001, ** p < 0.01 , * < 0.05. Abbreviations: CT, cortical thickness; GMV, gray matter volume; ICV, intracranial volume; n.s., not significant; SA, total SA; sGMV, subcortical GMV; WMV, white matter volume.

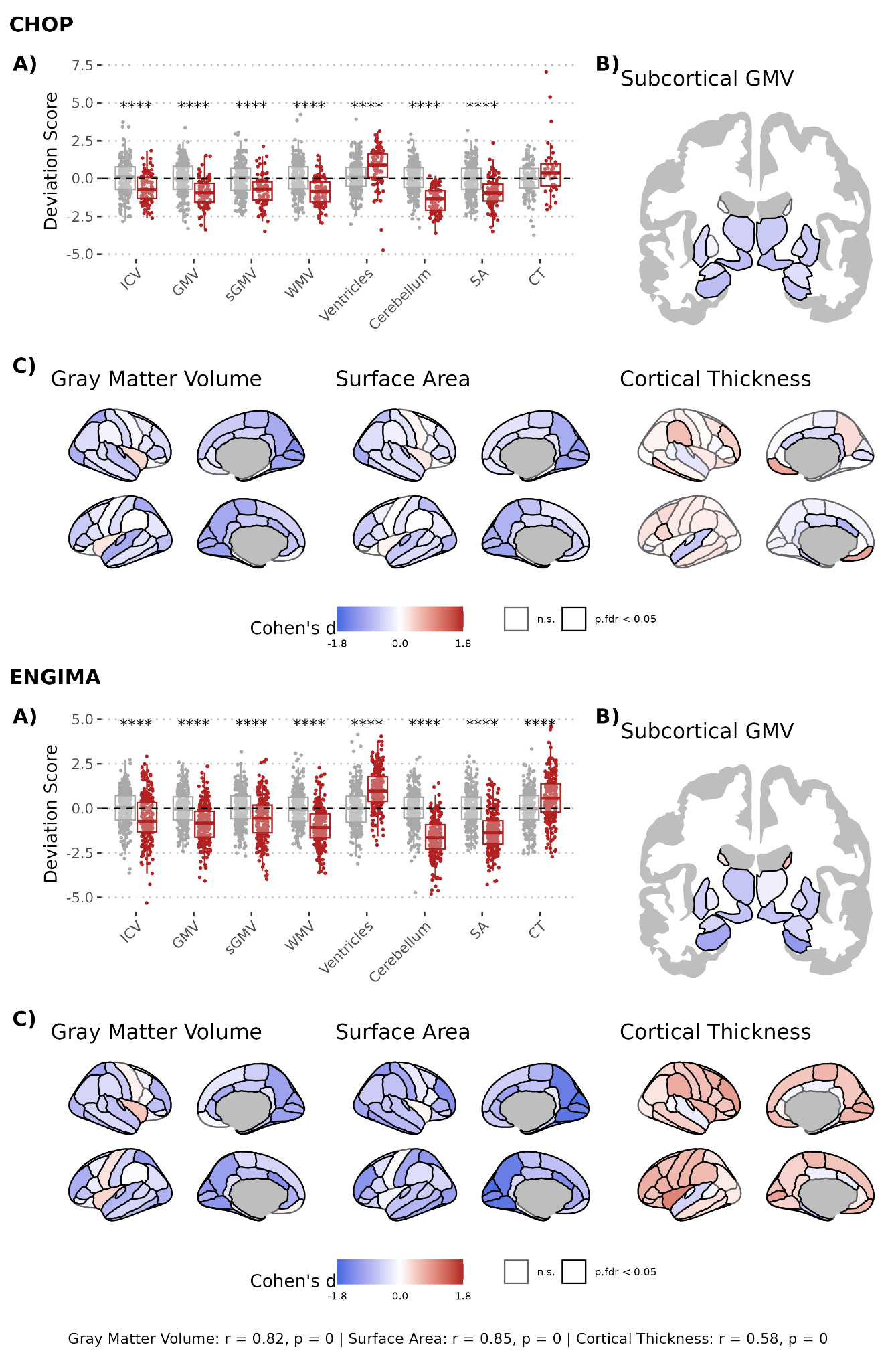

**Figure S18. Sensitivity analysis: Inclusion of fixed effect for scanner.** Cohen’s d effect sizes for the CHOP clinical primary cohort (left panel) and the ENIGMA-22q research cohort (right panel). Compared to the primary analysis, scanner was added as a covariate to the model to account for further scanner heterogeneity. (A) Box plot show individual standardized deviation scores for global brain measures in patients with 22q11DS (N CHOP = 92, N ENIGMA-22q = 242) and controls (N CHOP = 252, N ENIGMA-22q = 277). **(B and C)** Brain maps display Cohen’s d effect sizes for regional deviations in **(B)** subcortical gray matter volume (GMV) and **(C)** cortical GMV, surface area (SA), and thickness brain features. Regions with negative deviation scores in 22q11DS (i.e., smaller values in 22q11DS) are shown in blue, and regions with positive scores (i.e., larger values in 22q11DS) are shown in red. Solid black outlines indicate regions with significant case-control differences. **(D)** Scatter plots show the correlation between Cohen’s d effect sizes from the primary CHOP cohort (x-axis) and the independent ENIGMA-22q cohort (y-axis). All deviation scores were derived from normative growth charts predicting brain features based on age, sex, and scanner. P-values were corrected for multiple comparisons using the Benjamini-Hochberg false discovery rate (FDR) procedure. Asterisks indicate the level of statistical significance after correction. **** p < 0.0001, *** p < 0.001, ** p < 0.01 , * < 0.05. Abbreviations: CT, cortical thickness; GMV, gray matter volume; ICV, intracranial volume; n.s., not significant; SA, total SA; sGMV, subcortical GMV; WMV, white matter volume.

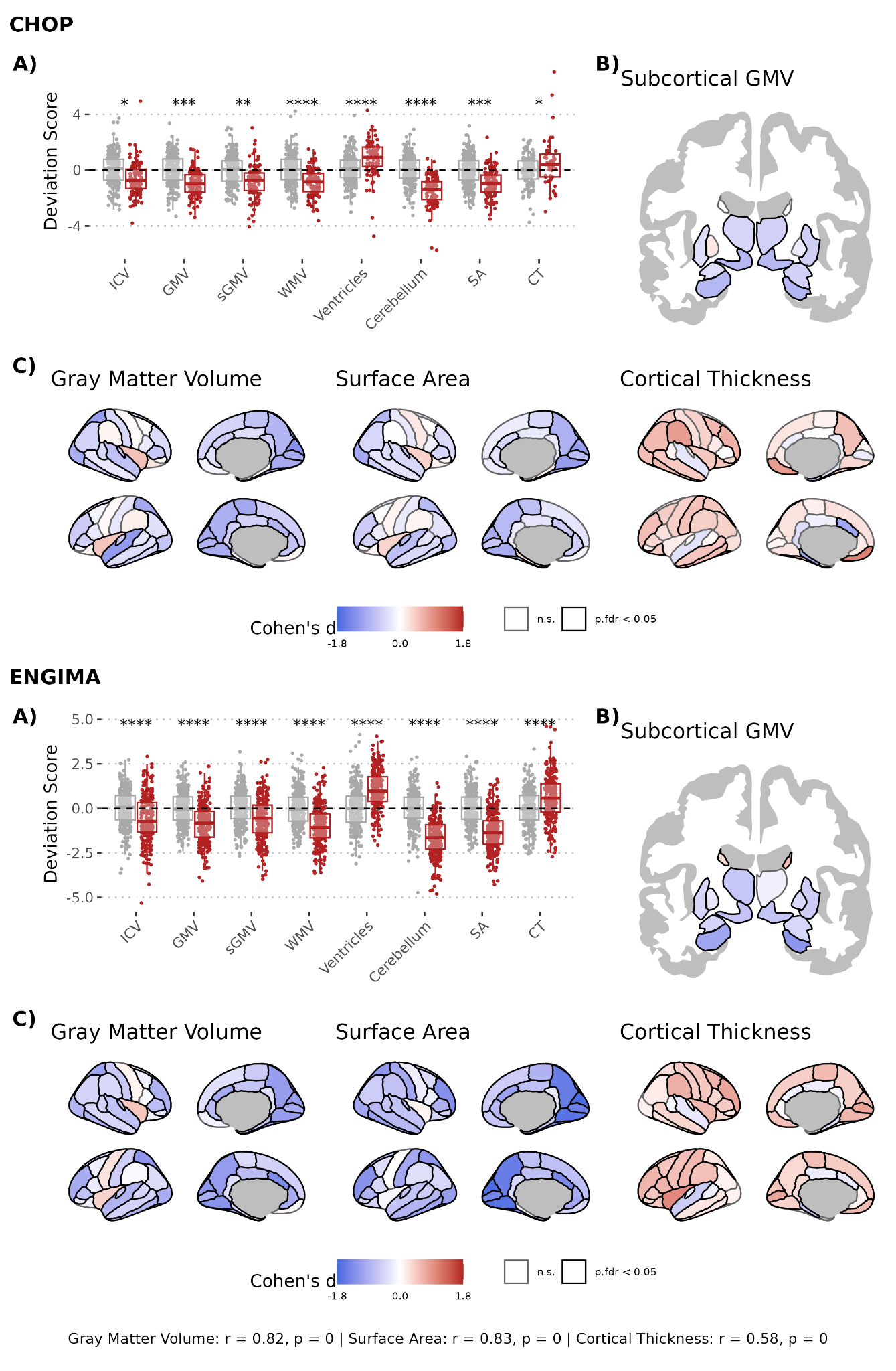

**Figure S19. Sensitivity analysis: Scans in participants older than 6 years.** Cohen’s d effect sizes for the CHOP clinical primary cohort (left panel) and the ENIGMA-22q research cohort (right panel). Compared to the primary analysis, we excluded brain measurements in patients younger than 6 years. (A) Box plot show individual standardized deviation scores for global brain measures in patients with 22q11DS (N CHOP = 55, N ENIGMA-22q = 242) and controls (N CHOP = 146, N ENIGMA-22q = 277). **(B and C)** Brain maps display Cohen’s d effect sizes for regional deviations in **(B)** subcortical gray matter volume (GMV) and **(C)** cortical GMV, surface area (SA), and thickness brain features. Regions with negative deviation scores in 22q11DS (i.e., smaller values in 22q11DS) are shown in blue, and regions with positive scores (i.e., larger values in 22q11DS) are shown in red. Solid black outlines indicate regions with significant case-control differences. **(D)** Scatter plots show the correlation between Cohen’s d effect sizes from the primary CHOP cohort (x-axis) and the independent ENIGMA-22q cohort (y-axis). All deviation scores were derived from normative growth charts predicting brain features based on age, sex, and scanner. P-values were corrected for multiple comparisons using the Benjamini-Hochberg false discovery rate (FDR) procedure. Asterisks indicate the level of statistical significance after correction. **** p < 0.0001, *** p < 0.001, ** p < 0.01 , * < 0.05. Abbreviations: CT, cortical thickness; GMV, gray matter volume; ICV, intracranial volume; n.s., not significant; SA, total SA; sGMV, subcortical GMV; WMV, white matter volume.

### References

1. Arcus Data Repository Team. Arcus Data Repository. Arcus at Children’s Hospital of Philadelphia; 2020.

2. Schabdach JM, Williams RMS, Logan J, Padmanabhan V, D’Aiello III R, Mclaughlin J, et al. From Scanner to Science: Reusing Clinically Acquired Medical Images for Research. AMIA Jt Summits Transl Sci Proc. 2025;2025:471–80.

3. Covitz S, Tapera TM, Adebimpe A, Alexander-Bloch AF, Bertolero MA, Feczko E, et al. Curation of BIDS (CuBIDS): A workflow and software package for streamlining reproducible curation of large BIDS datasets. NeuroImage. 2022;263:119609. https://doi.org/10.1016/j.neuroimage.2022.119609

4. Halchenko YO, Goncalves M, Ghosh S, Velasco P, Castello MV di O, Salo T, et al. HeuDiConv — flexible DICOM conversion into structured directory layouts [Internet]. Zenodo; 2025 [cited 2025 Aug 25]. https://doi.org/10.5281/zenodo.15080551

5. Morrow BE, McDonald-McGinn DM, Emanuel BS, Vermeesch JR, Scambler PJ. Molecular genetics of 22q11.2 deletion syndrome. Am J Med Genet A. 2018;176:2070–81. https://doi.org/10.1002/ajmg.a.40504

6. Zhou B, Purmann C, Guo H, Shin G, Huang Y, Pattni R, et al. Resolving the 22q11.2 deletion using CTLR-Seq reveals chromosomal rearrangement mechanisms and individual variance in breakpoints. Proceedings of the National Academy of Sciences. Proceedings of the National Academy of Sciences; 2024;121:e2322834121. https://doi.org/10.1073/pnas.2322834121

7. Iglesias JE, Billot B, Balbastre Y, Magdamo C, Arnold SE, Das S, et al. SynthSR: A public AI tool to turn heterogeneous clinical brain scans into high-resolution T1-weighted images for 3D morphometry. Sci Adv. 2023;9:eadd3607. https://doi.org/10.1126/sciadv.add3607

8. Billot B, Magdamo C, Cheng Y, Arnold SE, Das S, Iglesias JE. Robust machine learning segmentation for large-scale analysis of heterogeneous clinical brain MRI datasets. Proc Natl Acad Sci U S A. 2023;120:e2216399120. https://doi.org/10.1073/pnas.2216399120

9. Billot B, Greve DN, Puonti O, Thielscher A, Van Leemput K, Fischl B, et al. SynthSeg: Segmentation of brain MRI scans of any contrast and resolution without retraining. Med Image Anal. 2023;86:102789. https://doi.org/10.1016/j.media.2023.102789

10. Gopinath K, Greve DN, Das S, Arnold S, Magdamo C, Iglesias JE. Cortical analysis of heterogeneous clinical brain MRI scans for large-scale neuroimaging studies [Internet]. arXiv; 2023 [cited 2026 Jan 16]. https://doi.org/10.48550/arXiv.2305.01827

11. Zimmerman D, Mandal AS, Jung B, Buczek MJ, Schabdach JM, Karandikar S, et al. A systematic protocol to identify “clinical controls” for pediatric neuroimaging research from clinically acquired brain MRIs. bioRxiv. 2025;2025.06.25.661530. https://doi.org/10.1101/2025.06.25.661530

12. Schabdach JM, Schmitt JE, Sotardi S, Vossough A, Andronikou S, Roberts TP, et al. Brain Growth Charts for Quantitative Analysis of Pediatric Clinical Brain MRI Scans with Limited Imaging Pathology. Radiology. 2023;309:e230096. https://doi.org/10.1148/radiol.230096

13. Brant-Zawadzki M, Gillan GD, Nitz WR. MP RAGE: a three-dimensional, T1-weighted, gradient-echo sequence--initial experience in the brain. Radiology. 1992;182:769–75. https://doi.org/10.1148/radiology.182.3.1535892

14. Fischl B. FreeSurfer. Neuroimage. 2012;62:774–81. https://doi.org/10.1016/j.neuroimage.2012.01.021

15. Ching CRK, Gutman BA, Sun D, Villalon Reina J, Ragothaman A, Isaev D, et al. Mapping Subcortical Brain Alterations in 22q11.2 Deletion Syndrome: Effects of Deletion Size and Convergence With Idiopathic Neuropsychiatric Illness. Am J Psychiatry. 2020;177:589–600. https://doi.org/10.1176/appi.ajp.2019.19060583

16. Sun D, Ching CRK, Lin A, Forsyth JK, Kushan L, Vajdi A, et al. Large-scale mapping of cortical alterations in 22q11.2 deletion syndrome: Convergence with idiopathic psychosis and effects of deletion size. Mol Psychiatry. 2020;25:1822–34. https://doi.org/10.1038/s41380-018-0078-5

17. Bethlehem RAI, Seidlitz J, White SR, Vogel JW, Anderson KM, Adamson C, et al. Brain charts for the human lifespan. Nature. 2022;604:525–33. https://doi.org/10.1038/s41586-022-04554-y

18. Dorfschmidt L, White S, Gardner M, Bedford S, Ball G, Edwards AD, et al. Charting structural brain asymmetry across the human lifespan [Internet]. bioRxiv; 2025 [cited 2025 July 30]. p. 2025.07.21.665924. https://doi.org/10.1101/2025.07.21.665924

19. Rigby RA, Stasinopoulos MD, Heller GZ, De Bastiani F. Distributions for Modeling Location, Scale, and Shape: Using GAMLSS in R. 1st ed. Chapman and Hall/CRC; 2019.

20. Hawrylycz MJ, Lein ES, Guillozet-Bongaarts AL, Shen EH, Ng L, Miller JA, et al. An anatomically comprehensive atlas of the adult human brain transcriptome. Nature. 2012;489:391–9. https://doi.org/10.1038/nature11405

21. Arnatkevic̆iūtė A, Fulcher BD, Fornito A. A practical guide to linking brain-wide gene expression and neuroimaging data. NeuroImage. 2019;189:353–67. https://doi.org/10.1016/j.neuroimage.2019.01.011

22. Quackenbush J. Microarray data normalization and transformation. Nat Genet. 2002;32 Suppl:496–501. https://doi.org/10.1038/ng1032

23. Hawrylycz M, Miller JA, Menon V, Feng D, Dolbeare T, Guillozet-Bongaarts AL, et al. Canonical genetic signatures of the adult human brain. Nat Neurosci. 2015;18:1832–44. https://doi.org/10.1038/nn.4171

24. Fulcher BD, Little MA, Jones NS. Highly comparative time-series analysis: the empirical structure of time series and their methods. Journal of The Royal Society Interface. Royal Society; 2013;10:20130048. https://doi.org/10.1098/rsif.2013.0048

25. Seidlitz J, Mallard TT, Vogel JW, Lee YH, Warrier V, Ball G, et al. The molecular genetic landscape of human brain size variation. Cell Rep. 2023;42:113439. https://doi.org/10.1016/j.celrep.2023.113439

26. Dear R, Wagstyl K, Seidlitz J, Markello RD, Arnatkevičiūtė A, Anderson KM, et al. Cortical gene expression architecture links healthy neurodevelopment to the imaging, transcriptomics and genetics of autism and schizophrenia. Nat Neurosci. Nature Publishing Group; 2024;27:1075–86. https://doi.org/10.1038/s41593-024-01624-4

27. Low KJ, Foreman J, Hobson RJ, Kwuo H, Martinez-Cayuelas E, Almoguera B, et al. The LMSz method - an automatable scalable approach to constructing gene-specific growth charts in rare disorders. medRxiv. 2024;2024.08.19.24312213. https://doi.org/10.1101/2024.08.19.24312213

28. World Health Organization. WHO child growth standards: length/height-for-age, weight-for-age, weight-for-length, weight-for-height and body mass index-for-age: methods and development [Internet]. [cited 2025 Aug 25]. https://www.who.int/publications/i/item/924154693X. Accessed 25 Aug 2025
